# Covid-19 Risk by work-related factors: Pooled analysis of individual linked data from 14 cohorts

**DOI:** 10.1101/2023.12.19.23298502

**Authors:** Matthew Gittins, Jacques Wels, Sarah Rhodes, Bozena Wielgoszewska, Jingmin Zhu, Richard J Shaw, Olivia KL Hamilton, Evangelia Demou, Anna Stevenson, Ellena Badrick, Rebecca Rhead, S Vittal Katikireddi, George B. Ploubidis, Martie van Tongeren

**Author notes:** **Ethics** The UK LLC has ethical approval from the Health Research Authority Research Ethics Committee (Haydock Committee; ref: 20/NW/0446). **Data availability** The data is available on request from the UK-LLC https://ukllc.ac.uk/. The analysis code including detailing how variables were derived for each longitudinal cohort study are available from https://github.com/UKLLC/llc_0007. Understanding Society is an initiative funded by the Economic and Social Research Council and various Government Departments, with scientific leadership by the Institute for Social and Economic Research, University of Essex, and survey delivery by NatCen Social Research and Kantar Public. The Understanding Society COVID-19 study is funded by the Economic and Social Research Council (ES/K005146/1) and the Health Foundation (2076161). The research data are distributed by the UK Data Service. Generation Scotland received core support from the Chief Scientist Office of the Scottish Government Health Directorates [CZD/16/6] and the Scottish Funding Council [HR03006]. Genotyping of the GS:SFHS samples was carried out by the Genetics Core Laboratory at the Wellcome Trust Clinical Research Facility, Edinburgh, Scotland and was funded by the Medical Research Council UK and the Wellcome Trust (Wellcome Trust Strategic Award “STratifying Resilience and Depression Longitudinally” (STRADL) Reference 104036/Z/14/Z). Generation Scotland is funded by the Wellcome Trust (216767/Z/19/Z). Recruitment to this study was facilitated by SHARE - the Scottish Health Research Register and Biobank. SVK acknowledges funding from a NRS Senior Clinical Fellowship (SCAF/15/02). SVK acknowledge funding from the Medical Research Council (MC_UU_00022/2) and the Scottish Government Chief Scientist Office (SPHSU17). JW acknowledges funding from the Belgian National Scientific Fund (FNRS) CQ grant n°40010931. OKLH, RJS, SVK, and ED acknowledges funding from the Medical Research Council [MC_UU_00022/2] and the Scottish Chief Scientist Office [SPHSU17]. Note these codes will apply to all listed UoG authors. RJS is funded by Health Data Research UK (SS005). Role of funder. The funders had no role in the methodology, analysis or interpretation of the findings presented in this manuscript.

## Abstract

**Background:** SARS-CoV-2 infection rates vary by occupation, but the association with work-related characteristics (such as home working, key-worker, or furlough) are not fully understood and may depend on ascertainment approach. We assessed infection risks across work-related characteristics and compared findings using different ascertainment approaches.

**Methods:** Participants of 14 UK-based longitudinal cohort studies completed surveys before and during the COVID-19 pandemic about their health, work, and behaviour. These data were linked to NHS digital health records, including COVID-19 diagnostic testing, within the UK Longitudinal Linkage Collaboration (UK-LLC) research environment. Poisson regression modelled self-reported infection and diagnostic test confirmed infection within each cohort for work-related characteristics. Risk Ratios (RR) were then combined using random effects meta-analysis.

**Results:** Between March 2020 and March 2021, 72,290 individuals completed 167,302 surveys. Overall, 11% of 138,924 responses self-reported an infection, whereas 1.9% of 159,820 responses had a linked positive test. Self-reported infection risk was greater in key-workers vs not (RR=1.24(95%C.I.=1.17,1.31), among non-home working (1.08(0.98,1.19)) or some home working (1.08(0.97,1.17)) vs all home working. Part-time workers vs full-time (0.94(0.89,0.99)), and furlough vs not (0.97(0.88,1.01)) had reduced risk. Results for the linked positive test outcome were comparable in direction but greater in magnitude e.g. an 1.85(1.56,2.20) in key-workers.

**Conclusion:** The UK-LLC provides new opportunities for researchers to investigate risk factors, including occupational factors, for ill-health events in multiple largescale UK cohorts. Risk of SARS-CoV-2 infection and COVID-19 illness appeared to be associated with work-related characteristics. Associations using linked diagnostic test data appeared stronger than self-reported infection status.

**What is already known on this topic:**

- Infection of SARS-CoV2 during the pandemic was shown to vary by occupation, with occupations such as healthcare, and education at higher risk during some or all of the pandemic.
- What is not clear, is how are work-related characteristics such employment status, part-time working from home, and schemes such as furlough and key worker status associated with the risk of infection.

**What this study adds:**

- This is the one of the first studies to examine work-related characteristics including work related government policies, in terms of their infection risk within the working population.
- This is also one of the first studies to analyse data from the UK Longitudinal Linkage Collaboration (UK-LLC), in which multiple UK national longitudinal cohorts were linked to national health data including diagnostic testing for SARS-CoV2.
- We further compared definitions infection via either a self-reported case of COVID-19 or a linked diagnostic SARS-CoV2 infection.

**How this study might affect Research Practice or Policy:**

- The findings contribute to our understanding of work-related characteristics and related schemes were associated with infection risk under two definitions. This is pertinent given new and emerging variants are continuing to drive an ever-changing SARS-CoV-2 infection risk within the population, along with the need to adequately prepare for future pandemics that may occur.

## Introduction

Thought to be driven by the new BA.2.86 “Pirola” variant, between the 1^st^ July and 1^st^ October 2023 the estimated number of new cases SAR-CoV-2 infection, hospital admissions, and deaths with COVID have been steadily increasing^1 2^. In the UK, rates of SARS-CoV-2 infection and COVID-19-related mortality have been shown to vary by occupational group and have also changed over the course of the pandemic.^3-8^ Such variations could be related to differences in the working environment particularly exposure to other infected people, e.g. the ability to physically distance, indoor versus outdoor working, surface contacts^9-11^, differences in vaccination rate within the workforce, differences in infection mitigation strategies^14^ and the relaxation of these over time. These exposure risks are thought to be mitigated within the working population during and since the pandemic by characteristics of the work itself for example working from home. ^15 16^During the pandemic furlough and work from home orders were designed to restrict exposure risk, whilst simultaneously necessitating certain occupations as keyworkers with potential for an increased exposure. Studies investigating keyworker status and inability to work from home have tended to confirm these assumptions, ^17 18^ however a study of the ONS infection survey reported contradictory results for furlough than the expectation.^19^ Some of the restrictions imposed during lockdown have changed working habits over the long-term with a greater proportion of the population working from home or hybrid working.^20^ These working characteristics, including employment status itself, amount of part-time vs full time work, and the amount of time working from home are thought to be important characteristics in an employee’s risk of exposure. Other than key worker status and furlough, there is currently limited evidence to confirm if these measures did affect infection risk during the pandemic.

Ascertainment of SARS-CoV-2 infection can be via a test such as a PCR test or lateral flow test and previous SARS-CoV-2 infection can be identified via an antibody blood test. Infection status or history can also be ascertained via self-report – although this is contingent on individuals experiencing a symptomatic infection, otherwise known as COVID-19 illness. Observational studies exploring SARS-CoV-2 infection have tended to use self-reported COVID-19 illness through necessity to derive outcomes. Testing strategies in the UK varied during the pandemic and testing propensities have been linked to occupation – e.g., additional testing for NHS staff and schoolteachers at the start of the pandemic. Furthermore, the willingness to take a test may be linked to occupation.^21 22^ For instance, a healthcare worker might have had a higher propensity to take a test (due to fear of passing COVID-19 to vulnerable residents), compared to a farmer who works largely alone and outdoors. For this reason, studies that utilise test results (either self-reported or via health service records) are prone to bias. Use of the UK-LLC provides a unique opportunity to compare and combine different types of outcome data relating to SARS-CoV-2 infection.

During the pandemic a collaboration was formed between two UK National Core Studies (NCS), the Longitudinal Health and Wellbeing core study and the Transmission and Environment (aka PROTECT) core study. These initiatives brought together data from multiple UK population-based longitudinal studies to answer priority pandemic-related questions. As part of the Longitudinal Health and Wellbeing study,^23^ the UK Longitudinal Linkage Collaboration (UK LLC) was set up to bring together already established longitudinal studies with Electronic Health Records (EHR) from NHS Digital.^24^

### Aims

This study aimed to understand which work-related characteristics were associated with an risk of Covid-19 infection. We explored whether associations varied over time and according to differential ascertainment of infection status specifically self-reported infection vs diagnostic test defined infection. The work-related characteristics of interest available included: employment status (employed/full time/part-time/unemployed); keyworker status; furlough status; home working (fully/partially/not working from home).

## Methods

### Study Design & Data sources

Data for this study came from 14 population-based longitudinal cohort studies (see Supplementary Table S1, and S8) and linked electronic health records The UK Longitudinal Linkage Collaboration (UK LLC) is a Trusted Research Environment developed and operated by the Universities of Bristol and Edinburgh using an underlying ‘Secure eResearch Platform’ infrastructure (https://serp.ac.uk/) provided by Swansea University for longitudinal research. The UK LLC TRE is designed to host de-identified data from many interdisciplinary, longitudinal population studies; to systematically link these to participants’ health, administrative and environmental records; and to provide a secure analysis environment. This project has been approved by UK LLC and its contributing data owners and information on this project and its outputs can be accessed via UK LLC’s website (<u>Data Use Register | UK Longitudinal Linkage Collaboration</u> (ukllc.ac.uk) and UK LLC’s GitHub (<u>UK Longitudinal Linkage Collaboration GitHub</u>). Further details on each study can be found on the <u>UK-LLC documentation</u>. Where possible each study based in England was linked to Electronic Patient Health Records (EHR) available within the UK-LCC to provide diagnostic test infection data. Data forming the Covid-19 Second Generation Surveillance Systems contained Pillar 1 swab testing in PHE labs and NHS hospitals and Pillar 2 Swab testing in the community.^25^ The Covid-19 UK Non-hospital Antibody Testing Results (Pillar 3) was also linked.^26^ Additionally, vaccination data was sourced via the NHS COVID-19 Vaccination Status

### Work-related Exposure Variables

Individual longitudinal cohorts do not report information consistently, we therefore needed to harmonise across cohorts. The following were identified as suitably consistent work-related characteristics, self-reported within the longitudinal cohort studies:

- Economic status defined as unemployed, any employment/self-employment, and retired
- Employment status defined those employed into part-time and full time (either self-reported or when available working less than 35h a week).
- Home working (fully or partially) versus working at employer’s premises
- Keyworker status versus not keyworker status
- Furloughed versus not furloughed

### Covariates

Key demographic and socio-economic characteristics were defined as age group at the March 2020 (18-34; 35-44; 45-54; 55-66), sex (male/female/unknown), ethnicity (white/other), household composition (lives with partner and children, partner and no children, children only, alone, or other (e.g., housemates)), Urban/rural living. Linked SARS-CoV-2 vaccination status was obtained, where available, towards the end of the study period.

### Outcome Variables

Two binary (yes/no) dependent variables were investigated, Self-reported infection and diagnostic test confirmed infection as confirmed by a EHR linked positive test result (Pillar 1, 2, or 3). In each case this is defined as occurring since either the start of the pandemic or the previous within cohort survey (whichever is more recent). Self-reported infection was identified within each Longitudinal Cohort Study through either a self-reported infection when asked directly e.g., ‘do you think you have had COVID-19?’, or a self-reported positive test result.

### Statistical analysis

The analytic sample was first restricted to those of working age 18 to 66 years (inclusive) at the start of the pandemic. To appropriately investigate the work-related characteristics (Working from home, keyworker, and Furlough status) the analytical sample was further restricted to those self-reporting they were employed, self-employed (currently working or non-working), or furloughed (paid or unpaid leave from employment). Supplementary Material S2 and UK-LLC GitLab repository for further coding details. We investigated infection rates over the duration of the pandemic by stratifying the study-period into three time-periods (T1: April-June 2020, T2: July-October 2020, T3: November 2020-March 2021) to represent key periods of restrictions within the UK such as initial lockdown restrictions ending in June 2020 and the new lockdown and delta variant emerging in October 2020.^26 27^ Subsequently, time-periods T1 and T2, were combined due to limited numbers of events occurring in T2. The following analyses were repeated for the entire study period, and split by T1&2 and T3.^5^

We conducted a causal structure informed analysis using a previously published Direct Acyclic Graph (DAG)^28^ previously developed by the LHW and PROTECT teams for occupational related characteristics and COVID infection (Supplementary Figure S1).^5^ To account for the multilevel longitudinal structure associated with each cohort’s individual survey pattern, we fitted a mixed effects Poisson regression with robust standard errors and an individual level random intercept. The resulting risk ratios aided interpretation and avoided issues related to non-collapsibility of odds ratios.^29 30^ Each model included a set of predefined fixed effect covariates to investigate the influence of confounding whilst accounting for differential data availability across cohorts. We estimated unadjusted associations, and two adjustment models (Adj-Model 1 and Adj-Model 2). Model 1 included age group at the start of the pandemic, sex, and ethnicity. Then where studies allowed, model 2 adjusted for household composition, home location (rural vs urban), and SARS-CoV-2 vaccination status. Vaccination status was allowed to vary depending on the time period and was defined as ‘at least 1 dose’ versus none, as access to vaccination in the working age population was limited during the study period. Results cohort were pooled using a random effects meta-analysis with restricted maximum likelihood. Heterogeneity between studies was assess via appropriate statistics. All analyses were conducted in Stata 17.

## Results

Across all studies, 9.1% of responses to a within cohort survey indicated a self-reported COVID infection, whereas 1.8% were linked positive test result (Table 1 and Supplementary Table S3). Note, cohorts with a greater number of surveys performed during the time period (e.g. UKHLS) report a lower self-reported response rate than the majority of cohorts that performed 1 or 2 surveys during the period.

**Table 1.**
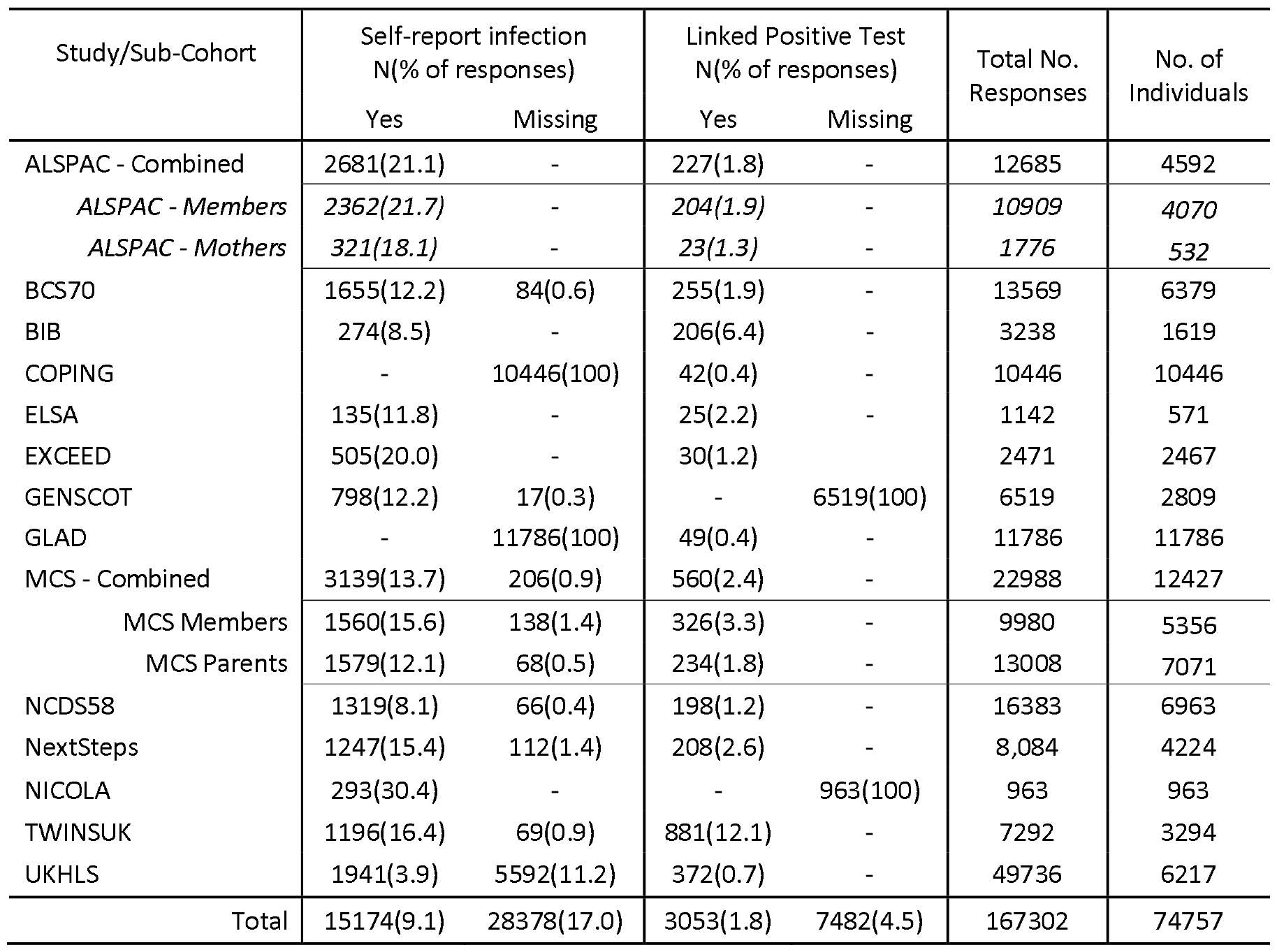
Self-reported infection, and EHR linked positive test response rates by longitudinal cohort study and sub-study.

Of the 15,174 self-reported infections 31% also reported a self-reported a positive infection test (see Supplementary Table S4), however only 9.6% also had a linked positive test. Excluding those with a missing self-reported response, 11% of 138,924 completed responses to waves reported a self-reported infection, 1.9% of 159,820 that could be linked to English EHR data had a linked positive test between April 2020 and March 2021. The within time-period infection rate defined from self-reported and linked diagnostic test increased for each additional time-period (see Supplementary Table S5) within the study. For each self-reported work-related characteristic, Table 2 contains infection rates per time-period and combined across the entire analysis study-period (Mar20-Mar21). Note, Supplementary Table S5 reports the equivalent information for covariate demographic statistics where available. For self-reported infection, those employed or looking for work (11% of responses with COVID), in full-time work (10.7%), key-worker status (13%), spent no time working from home (10.3%), and not in furlough (10.7%) all had a higher percentage of responses with self-reported infection compared to their comparison groups. Despite the smaller rates of self-reported infection & linked positive tests, the difference in rates were similar. Rates related to keyworker (2.5%) vs not (1.6%), Furlough (1.1%) vs Not (1.4%), and home working in particular ‘all the time’ (1.1%), ‘some of the time’ (1.5%), and ‘none of the time’ (2.1%) all appear to indicate consistent absolute differences with the self-reported infection.

**Table 2.** Self-reported and linked infection rates for work-related exposure characteristics by time-tranche, and full study period.

| Time-period | Time-period 1<br>March 2020 - October 2020 |  |  |  | Time Tranche-2<br>Nov 2020 - March 2021 |  |  |  | Full period<br>March 2020 - March 2021<br>(inc NICOLA) |  |  |  |
| --- | --- | --- | --- | --- | --- | --- | --- | --- | --- | --- | --- | --- |
| COVID Test-definition | Self-reported |  | Linked Test Result |  | Self-reported |  | Linked Test Result |  | Self-reported |  | Linked Test Result |  |
| Responses | %Pos | Total responses | %Pos | Total responses | %Pos | Total responses | %Pos | Total responses | %Pos | Total responses | %Pos | Total responses |
| Economic Status |  |  |  |  |  |  |  |  |  |  |  |  |
| Eco Active(Emp/looking) | 9.8 | 63,204 | 1.1 | 78,059 | 13.4 | 34,136 | 3.5 | 33,098 | 11.1 | 97,340 | 1.8 | 111,157 |
| Retired | 7.1 | 5,581 | 0.8 | 7,884 | 9.6 | 2,847 | 2.1 | 2,483 | 8 | 8,428 | 1.1 | 10,367 |
| Unemployed | 7.5 | 16,435 | 1.2 | 17,329 | 14 | 10,314 | 2.7 | 10,177 | 10 | 26,749 | 1.8 | 27,506 |
| Total | 9.2 | 85,220 | 1.1 | 103,272 | 13.3 | 47,297 | 3.2 | 45,758 | 10.6 | 132,517 | 1.7 | 149,030 |
| Employment Status |  |  |  |  |  |  |  |  |  |  |  |  |
| In Full-time Employment | 9.4 | 39,196 | 0.8 | 46,459 | 13 | 23,158 | 3.1 | 22,227 | 10.7 | 62,354 | 1.6 | 68,686 |
| In Part-time Employment | 8.1 | 12,232 | 0.6 | 20,197 | 13.8 | 6,298 | 3 | 6,309 | 10 | 18,530 | 1.2 | 26,506 |
| Retired | 6.1 | 4,371 | 0.2 | 6,564 | 9.5 | 2,701 | 1.6 | 2,336 | 7.4 | 7,072 | 0.6 | 8,900 |
| Employed but not working | 8.8 | 10,431 | 0.6 | 10,144 | 15 | 4,635 | 3.6 | 4,506 | 10.7 | 15,066 | 1.5 | 14,650 |
| Unemployed | 6.3 | 12,062 | 0.7 | 13,013 | 13.9 | 9,880 | 2.6 | 9,791 | 9.7 | 21,942 | 1.5 | 22,804 |
| Total | 8.4 | 78,292 | 0.7 | 96,377 | 13.3 | 46,672 | 2.9 | 45,169 | 10.2 | 124,964 | 1.4 | 141,546 |
| Key Worker |  |  |  |  |  |  |  |  |  |  |  |  |
| No | 10.1 | 25,482 | 1 | 28,522 | 12 | 11,697 | 2.8 | 11,763 | 10.7 | 37,179 | 1.6 | 40,285 |
| Yes | 12 | 24,420 | 1.6 | 28,165 | 15.1 | 13,991 | 4.3 | 14,067 | 13.2 | 38,411 | 2.5 | 42,232 |
| Total | 11.1 | 49,902 | 1.3 | 56,687 | 13.7 | 25,688 | 3.6 | 25,830 | 12 | 75,590 | 2 | 82,517 |
| Home Working |  |  |  |  |  |  |  |  |  |  |  |  |
| All time at home | 7.8 | 17,147 | 0.3 | 15,708 | 12.3 | 10,753 | 2.2 | 10,474 | 9.5 | 27,900 | 1.1 | 26,182 |
| Some time at home | 6.8 | 6,520 | 0.3 | 6,297 | 12.7 | 5,774 | 2.8 | 5,619 | 9.6 | 12,294 | 1.5 | 11,916 |
| No time at home | 7.9 | 20,639 | 1 | 19,604 | 13.8 | 13,716 | 3.7 | 13,247 | 10.3 | 34,355 | 2.1 | 32,851 |
| Total | 7.7 | 44,306 | 0.6 | 41,609 | 13.1 | 30,243 | 3 | 29,340 | 9.9 | 74,549 | 1.6 | 70,949 |
| Furlough |  |  |  |  |  |  |  |  |  |  |  |  |
| No | 9.4 | 45,906 | 0.6 | 61,556 | 12.9 | 27,829 | 3.2 | 27,118 | 10.7 | 73,735 | 1.4 | 88,674 |
| Yes | 8.7 | 7,122 | 0.4 | 7,505 | 15.6 | 3,120 | 2.8 | 2,987 | 10.8 | 10,242 | 1.1 | 10,492 |
| Total | 9.3 | 53,028 | 0.6 | 69,061 | 13.1 | 30,949 | 3.1 | 30,105 | 10.7 | 83,977 | 1.4 | 99,166 |

### Results: Work-related status within those employed or economically active (Home-working, key-worker, furlough)

Figure 1 reports the relative risk (RR) and 95% confidence intervals (95% C.I) obtained in stage two of the IPD meta-analysis. Each figure reports results (crude, Adj-Mod1, Adj-Mod2) by time-period and overall, for self-reported and linked positive outcomes.

**Figure 1.**
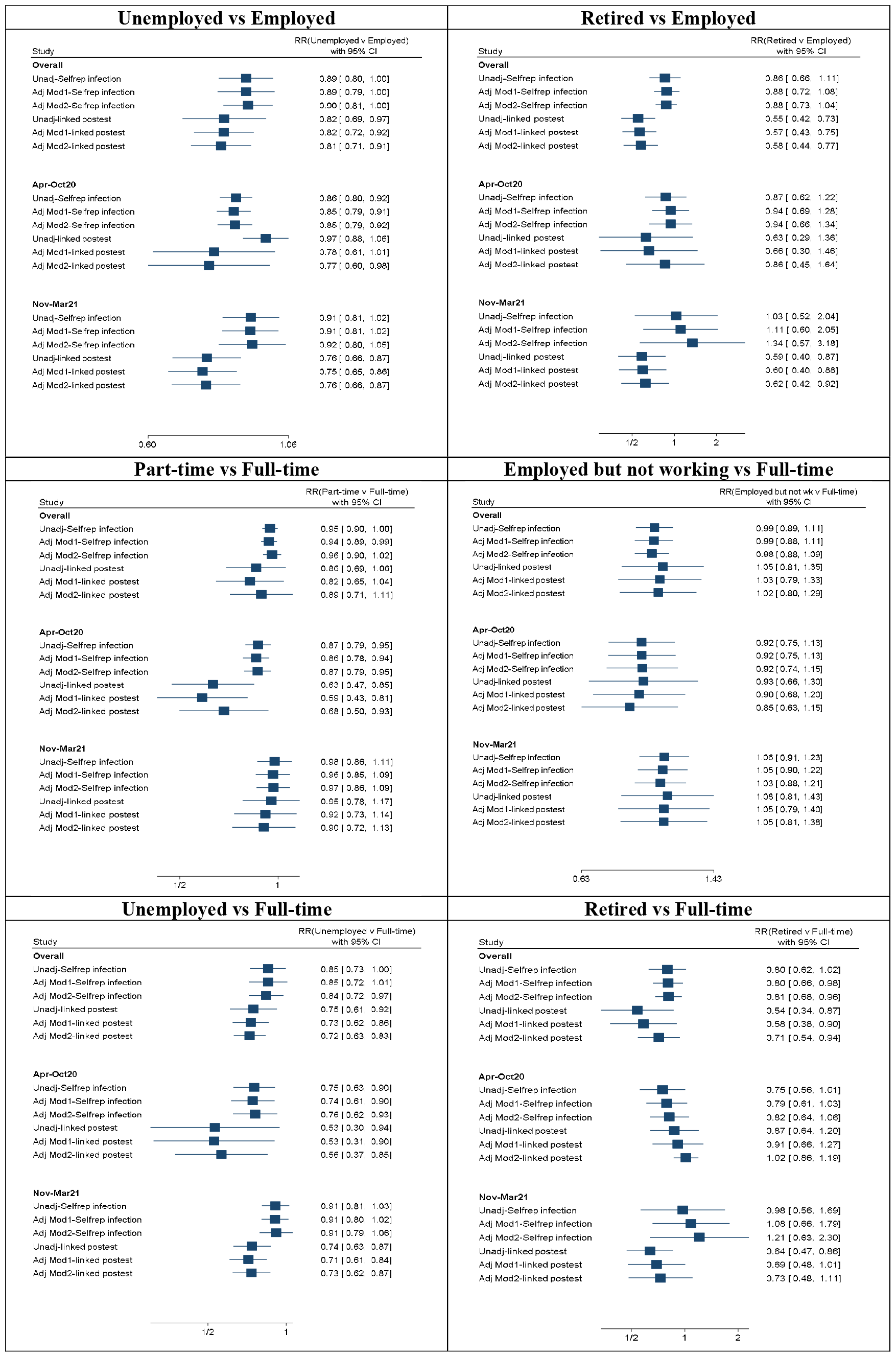
Two-stage IPD-meta analysis combined Relative Risk (95% C.I.) comparing Work-related Status for unadjusted and adjusted models of self-reported infection and linked positive infection by time-period. *Footnote to table: Adj Mod 1 includes age, gender and ethnicity, Adj-mod2 also includes where available* household composition, home location, and vaccination status.

Each panel within the figure reports results associated with the work-related characteristic and its reference category. Figure 1 includes investigated economic/employment status in the full sample under the two definitions 1. compare ‘unemployed’ and ‘retired’ against ‘employed/economically active’ and 2. compare ‘part-time’ employment, ‘employed but not working’, ‘unemployed’, ‘retired’ against ‘full time working’. These were followed by the models looking specifically at COVID 19-related work characteristics in those that were reported to be employed/economically active. Figure 2 repeats the results for 1. ‘some’ and ‘none’ home working vs ‘all’ home working, 2. key worker status yes v no, and 3. furlough status yes v no.

**Figure 2.**
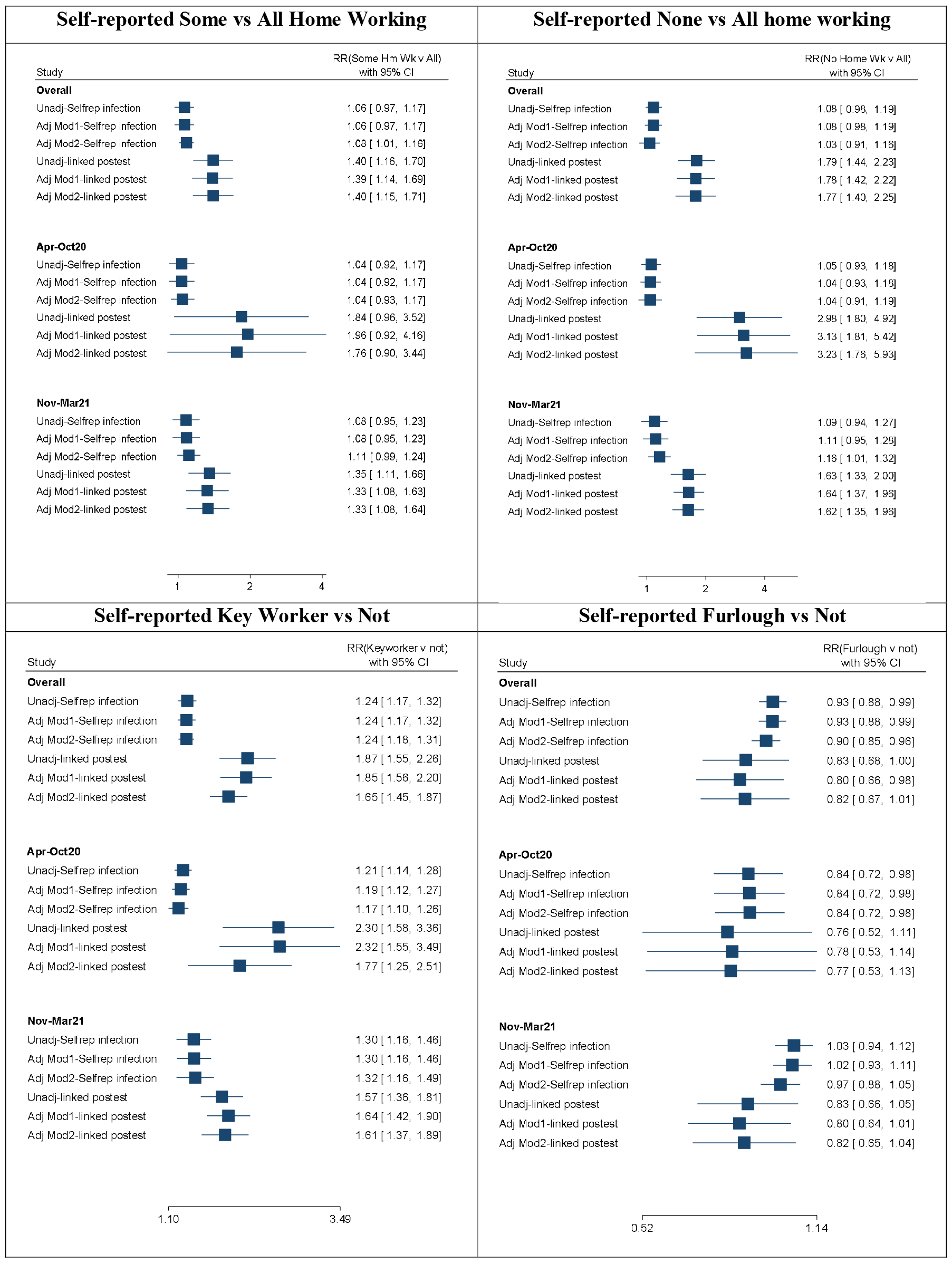
Results of two-stage IPD-meta analysis combined Relative Risk (95% C.I.) comparing work-related characteristics for unadjusted and adjusted models of self-reported infection and linked positive infection by time-period. Footnote to table: Adj Mod 1 includes age, gender and ethnicity, Adj-mod2 also includes where available household composition, home location, and vaccination status.

The results of contrasts between retirement and unemployment are reported but should be noted as highly unstable due to small within-study samples. Those in part-time employment were at 6% (RR(95%C.I) = 0.94(0.89,0.99)) and approximately 18% decreased risk (0.82(0.65,1.04)) of self-reported infection and linked COVID-19 positive test result. These associations both appear to be stronger in the first time-period April to October 2020, but tended towards the null between November and March 2021. Employed but not working compared to employed full-time showed evidence of a small decreased risk in the time-period T1&2, but this was not apparent in the time-period (T3).

### Results: Work-related characteristics within those employed or economically active (Homeworking, Key-worker, furlough)

‘Some’ home-working versus all home working indicated an increased relative risk of approximately 6% (RR=1.06, 95% C.I. = 0.97;1.17) in self-reported infection. This was slightly lower between Apr-Oct 2020 at 4% and slightly higher between Nov 2020 and March 2021 at 8%. Linked positive test at 39% indicated a larger increased RR compared to self-reported infection (RR=1.39 (1.14;1.69)). This was also greater between Apr-Oct 2020 at 96% (RR=1.96 (0.92;4.16) compared to Nov 2020 – Mar 2021 at 33%.

No home-working indicated an increased risk of approximately 8% (RR=1.08, 95% C.I. = 0.98;1.19) in self-reported infection compared to ‘all’ home working. This was slightly lower between Apr-Oct 2020 at 4% and slightly higher between Nov 2020 and March 2021 at 11%. Linked positive test indicated a larger RR compared to self-reported infection of 78% (RR=1.78 (1.42;2.22)). This was greater between Apr-Oct 2020 with RR=3.13 (1.81;5.42) than Nov 2020 – Mar 2021 at RR=1.64 (1.37;1.96).

Key-worker status indicated an increased risk of 24% (RR=1.24, 95% C.I. = 1.17;1.32) in self-reported infection compared to non-key workers. Linked positive test indicated a larger increased RR compared to self-reported infection of 85% (RR=1.85 (1.56;2.20)) for key-workers versus not. Furlough status indicated a decreased risk for those on furlough of approximately 7% (RR=0.93, 95% C.I. = 0.88;0.99) in self-reported infection. Linked positive test indicated a larger decreased RR compared to self-reported infection of 20% for furlough versus not. Again, in each case the RR were slightly lower between April-Oct 2020 and higher Nov 2020 – April 2021. Full results for all models including heterogeneity statistics, and example Forest plot for furlough status, can be found in Table S6, and Figure S7.

## Discussion/Conclusion

We observed that keyworkers, and those working some or all of their time away from the home, were associated with an increased risk of COVID infection. Whereas those that were furloughed, those retired, unemployed, or those working part-time rather than full-time were at decreased risk of infection. Differences in risk were greater during the first six months of the pandemic (Apr-Oct 2020) before dissipating as restrictions were lifted in the second six months (Nov20-Mar21). Though the direction of associations was unchanged throughout, their magnitude was greater when using a diagnostic test result as an indicator of COVID infection rather than self-reported infection.

Previous work looking at infection risk due to work-related characteristics has largely focused on the risk observed due to occupational groups, and how their risk changed over time.^4 5 12^ Here we have looked to understand work-related characteristics rather than occupation. Some of these characteristics were defined by government policies during the pandemic designed to reduce social interaction at work and thus reduce the spread of infection at the population level. Policies encouraging those who could work from home to do so, and the development of the furlough scheme, appear to have been successful in reducing risk. For self-reported infection those furloughed indicated a reduced risk by approximately 7% and for linked diagnostic positive test by 20%. These results appear contradicted a recent publication using the ONS infection survey that reported an 80% increased odds of COVID infection in furloughed individuals vs employed.^19^. These counter intuitive results, not replicated here, were thought explained by increased social interactions outside of work. As these initiatives were reduced over time, their effect also appeared to dissipate with self-reported infection returning to the null effect for furlough. Similarly key-workers, those occupations deemed essential and largely public facing such as health care and food retail, were observed to have an increased risk of infection. Interestingly here self-reported risk was slightly greater in the second time-period (30% increase up from 19%), but reduced for diagnostic test (64% increase down from 132% increase). This may have been due to changes in the definition of key workers over time, and increased access to diagnostic testing in non-key worker occupations.

It is likely that the occupation of the participant will have been a driving factor behind these results something we were unable to adequately account for. Those essential occupations requiring work to be performed away from the home such as healthcare, education, social care, and food production would have been important here due to their inability to work from home, their increased exposure, and their earlier and more regular access to diagnostic testing. Previous work, including those using robust testing procedures such as the ONS infection survey,^5 6 8^ also indicated that health-care workers were at increased risk during this time, and that risk was greatest within the equivalent April to October 2020 T1&2 before dissipating in the subsequent October 2020 to April 2021 equivalent to T3. Additionally Social and Education occupations, working from home during T1&2 had lower risk, but returning to the classroom in T3 resulted in an increased risk.

Using a diagnostic test rather than self-report to define infection consistently indicated greater relative risks for the same comparison. If differential access to diagnostic testing were present then we may expect biased relative risks for the diagnostic test outcome, but also self-reported infection as they would be able to confirm or refute suspected cases. Greater diagnostic testing in certain occupational groups during the early period of the pandemic may have driven overestimated effects due to diagnostic test availability. For example, healthcare workers had greater access to diagnostic testing than the general population, health care workers were also full-time and key workers, both of which showed elevated risk compared to the comparison group which didn’t have the same access to diagnostic testing hence lower rates in comparison the self-reported results. This is consistent with the corresponding effect for furlough workers, as here the non-furlough group would be expected to have greater access, meaning greater reduced effects though wide confidence intervals do include the effects for self-reported COVID-19. As was confirmed, as access to testing increased over time, we would expect these differences to also diminish. Unfortunately, we did not have access to negative test results, or to those testing positive through home testing only i.e. not seeking to confirm their positive result via an NHS testing centre and so were not able to investigate further.

### Strengths and Limitations of the study

The UK-LLC provides a unique opportunity to model infection risk in a large cohort of individuals across the UK and Ireland. This increased the sample size, and improved the representativeness of the combined cohort over a single cohort study. The use of multiple surveys across the pandemic and their linkage to the diagnostic testing and allowed for a more detailed comparison of results associated with self-reported infection and positive diagnostic testing. This meant for the first time we were able to compare and contrast results obtained under the two definitions of an infection.

However, there were a number of complex challenges. The data is observational in nature with largely self-reported information on the work-related factors and participant characteristics. Significant differences between cohort studies were present in the sampling methods, data collection, survey questions, and missing data. This and the subsequent harmonising process across studies resulted in a reduced ability to adopt more sophisticated single IPD modelling approach, employ sampling weights, and account for sources of confounding in suitable detail. We were not able to suitably investigate or account for occupation, and corresponding job characteristics relating to exposure risk via a Job Exposure Matrix.^10^ Additional factors not related to work such as physical contacts outside work, use of public transport, shopping, hospitality (restaurants, cafes, pubs), and occupations of family / cohabitants were either not available in suitable number of studies to be included. Small amounts of bias, particularly affecting marginalised groups, may have been introduced due to incomplete or incorrect matching, people not consenting to having their survey records linked to health records.^31^ Due to the self-reported cross-sectional nature of the surveys, including reporting infection since previous survey (or start of pandemic), we cannot infer the direction of causation. Additionally, the linked diagnostic testing data did not include linked data for Scotland and Ireland, and did not include negative test results and so a more optimal test-negative design was not possible here.^32^

## Conclusion

Policies aimed at reducing social interaction such as working from home, the furlough scheme were associated with reduced risk, whereas keyworkers most of whom were public facing were associated with increased risk. Since the pandemic began the Infection rates, prevalence, hospital admissions, and deaths with COVID continue to be in constant change.^1 2^ New and emerging variants, along with changes in population behaviour, drive these changes but also influence the ability of vaccines to provide resistance.^33 34^ This changing nature and the potential for future pandemics, mean working characteristics and mitigation strategies are still important considerations for policy makers. Future strategies should renew focus on reducing the infection risk posed by those unable to work from home including key worker status. Strategies such as improved supply of protective equipment, stronger enforcement of controls within the workplace, and better testing should be considered a priority. The definition of the infection (diagnostic testing v self-report) is an important consideration when interpreting studies that investigate infection risk using a single definition of infection, especially of the testing method is not readily available for all.

## Data Availability

The data is available on request from the UK-LLC https://ukllc.ac.uk/. The analysis code including detailing how variables were derived for each longitudinal cohort study are available from https://github.com/UKLLC/llc_0007.

https://ukllc.ac.uk/

https://github.com/UKLLC/llc_0007

## Acknowledgements

‘The UK LLC is an initiative of the UKRI-funded Longitudinal Health and Wellbeing National Core Study led by University College London (Grant code: MC_PC_20059). This work uses data accessed within UK LLC’s Trusted Research Environment (TRE), hosted by the Secure eResearch Platform (SeRP UK). We thank the SeRP UK Team at Swansea University and NHS Digital Health and Care Wales for providing the TRE’s infrastructure and support.

This work uses data provided by participants of the contributing LPS within the UK LLC TRE, which have been collected through their longitudinal study or as part of their care and support and/or interactions with UK government services. We wish to recognise and thank the study participants and each contributing LPS team, including data managers, administrators and those collecting data. We thank the following LPS for contributing data that made this research possible

- Avon Longitudinal Study of Parents and Children (ALSPAC)
- 1970 British Cohort Study (BCS70)
- Born in Bradford (BIB)
- English Longitudinal Study of Ageing (ELSA)
- Extended Cohort for E-health, Environment and DNA (EXCEED)
- Generation Scotland
- Genetic Links to Anxiety and Depression Study (GLAD)
- Millennium Cohort Study (MCS)
- National Child Development Study (NCDS)
- Psychiatry and Neurological Genetics (COPING) Study
- Next Steps
- Northern Ireland Cohort for the Longitudinal Study of Ageing (NICOLA)
- TwinsUK
- The UK Household Longitudinal Study (Understanding Society).

A full list of acknowledgments, including support for each study, is provided in the supplementary materials. We thank the NHS and particularly NHS Digital for their work in curating participants’ health records and for making these available for public benefit research designed to improve health services.

