## Supplementary Material for "Covid-19 Risk by work-related factors: Pooled analysis of individual linked data from 14 cohorts"

#### S1. Supplementary Material – Summary statistics

***Figure S1 - Directed Acyclic Graph of hypothesised causal structure.***

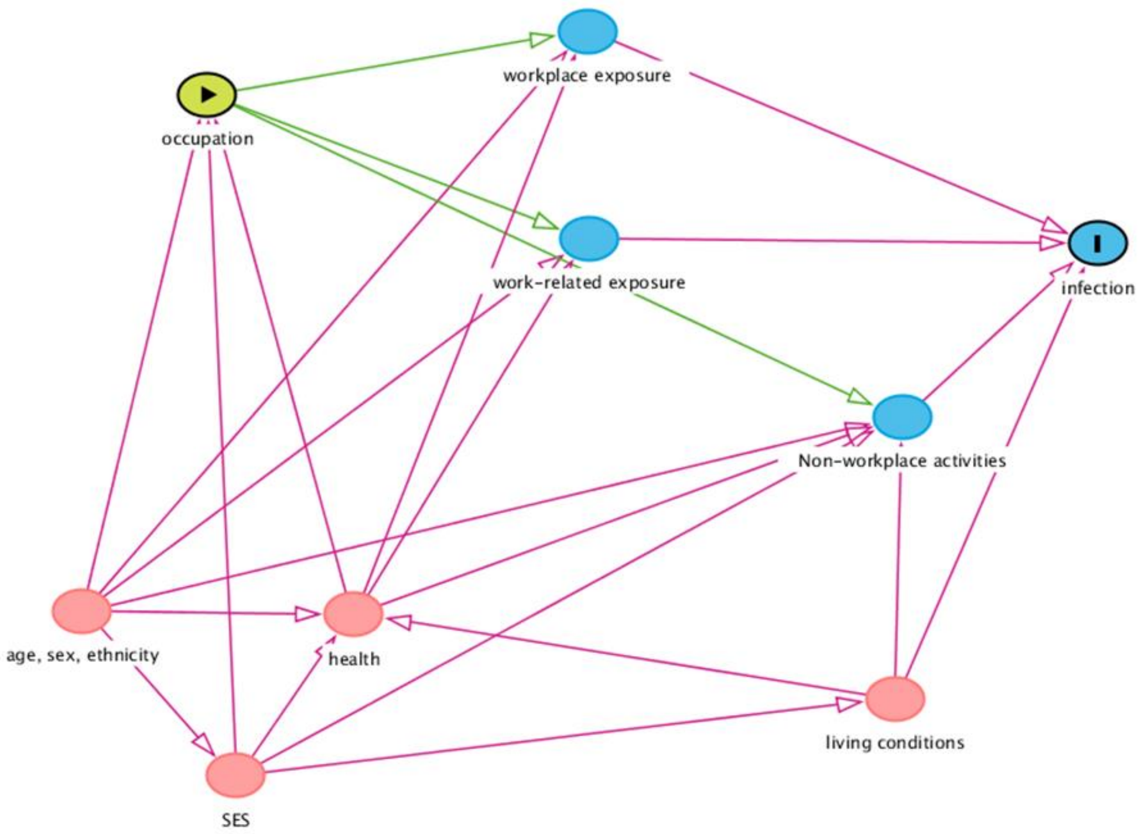

*Table S1 - Summary of the longitudinal cohort studies within the UK-LLC*

| Study ID | Full Study Name | Description | First Survey | Last Survey | Number of pandemicSurvey Waves used |
|---|---|---|---|---|---|
| ALSPAC | Avon Longitudinal Study of Parents and Children | The children (and some of their mothers) of women who were pregnant between Apr 1991 Dec 1992 in South West England.[18 19] | Apr-May 20 | Nov20-Mar21 | 4 |
| BCS70* | British Cohort Study 1970 | Children born in the UK in one week in 1970, with regular follow-up surveys from birth.[20] | May-20 | Feb/ Mar-21 | 3 |
| BIB | Born in Bradford | Pregnant women (& partners) recruited at Royal Bradford Infirmary between 2007-2011.[21 22] | Apr-Jun 20 | Oct-Dec 20 | 2 |
| COPING | COVID-19 Psychiatry and Neurological Genetics Study | Aged 16+ recruited from existing cohorts hosted by NIHR BioResource (GLAD, the Eating Disorders Genetics Initiative (EDGI)).[23] | Apr-20 | Apr-20 | 1 |
| ELSA | English Longitudinal Study of Ageing | Nationally representative pop aged 50+, in private households in England, includes biennial surveys, periodic refreshing of sample. [24] | June/Jul 2020 | Nov/Dec 2020 | 2 |
| EXCEED | Extended Cohort for E-health, Environment and DNA | Cohort aged 40-69 recruited 2013+, from the general population through general practices in Leicestershire and Rutland.[25 26] | Apr 21 | Apr 21 | 1 |
| GENSCOT | Generation Scotland | A family-based genetic epidemiology study of volunteers from across Scotland, aged 18+.[27] | May-20 | Mar-21 | 3 |
| GLAD | Genetic Links to Anxiety and Depression | Participants with depression and/or anxiety aged 16+, invited via media campaigns and recruited online into the NIHR BioResource.[28] | Apr-20 | Apr-20 | 1 |
| MCS* | Millennium Cohort Study | UK children born Sep 2000 - Jan 2002, with regular follow-up surveys from birth. [29] | May-20 | Feb/Mar-21 | 3 |
| NCDS58* | National Child Development Study 1958 | UK Children born in one week in 1958, with regular follow-up surveys from birth.[30] | May-20 | Feb/Mar-21 | 3 |
| NextSteps* | Next Steps 1989 | Recruited via secondary schools in England at age 13, regular follow-up surveys thereafter.[31] | May-20 | Feb/Mar-21 | 3 |
| NICOLA | Northern Ireland Cohort for the Longitudinal Study of Ageing | aged 50+, recruited 2012+. Randomized sample of NI addresses from Business Service Organization (BSO) GP Register Database.[32] | Feb-Mar 20 | Feb-Mar 20 | 1 |
| TWINSUK | Twins UK study | Volunteer adult twins (55% monozygotic/43% dizygotic) from UK aged 18+.[34] | Apr-May 20 | Oct-Nov 20 | 3 |
| UKHLS | The UK Household Longitudinal Study (Understanding Society) | A nationally representative longitudinal household panel survey, based on a clustered-stratified probability sample of UK households, with participants surveyed annually.[35] | Apr-20 | Mar-21 | 8 |

*Age homogenous cohorts: participants all born in the same year or week of the same year

*Table S2 - List of the models tested in the study with exposure modalities and sample subsets.*

| Model | Exposure | Modalities | Sample subset |
|---|---|---|---|
| 1 | Unemployed | (1) Unemployed | All respondents aged 18-66 |
| | | (0) Employed | |
| | Retired | (1) Retired | |
| | | (0) Employed | |
| 2 | Part-time | (1) Part-time | |
| | | (2) Full-time | |
| | Employed not working | (1) Employed but not working | |
| | | (0) Full-time | |
| | Unemployed | (1) Unemployed | |
| | | (0) Full-time | |
| | Retired | (1) Retired | |
| | | (0) Full-time | |
| 3 | Key worker | (1) key worker | Respondents employed aged 18-66 i.e. self-defined as employed, self-employed, or looking for work |
| | | (0) Not key worker | |
| 4 | Partial home working | (1) Partial home working | |
| | | (0) no home working | |
| | No home working | (1) Full-time home working | |
| | | (0) no home working | |
| 5 | Furlough | (1) Furlough | |
| | | (0) Not Furlough | |

*Table S3 – COVID-19 Infection summary statistics by cohort study*

| Study/Sub-Cohort | Self-report infection | | Self-report Positive Test Only | | Linked Positive Test | | Total Responses | No Individuals |
|---|---|---|---|---|---|---|---|---|
| | Yes | Missing | Yes | Missing | Yes | Missing | | |
| ALSPAC - Combined | 2681(21.1) | - | 695(5.5) | - | 227(1.8) | - | 12,685 | 4592 |
| ALSPAC - Members | 2362(21.7) | - | 629(5.8) | - | 204(1.9) | - | 10,909 | |
| ALSPAC - Mothers | 321(18.1) | - | 53(3.0) | - | 23(1.3) | - | 1,776 | |
| BCS70 | 1655(12.2) | 84(0.6) | 445(3.3) | 84(0.6) | 255(1.9) | - | 13,569 | 6379 |
| BIB | 274(8.5) | - | 92(2.84) | - | 206(6.4) | - | 3,238 | 1619 |
| COPING | - | 10446(100) | - | 10446(100) | 42(0.4) | - | 10,446 | 10446 |
| ELSA | 135(11.8) | - | 23(2.0) | - | 25(2.2) | - | 1,142 | 571 |
| EXCEED | 505(20.0) | - | 48(1.9) | | 30(1.2) | | 2,471 | 2462 |
| GENSCOT | 798(12.2) | 17(0.3) | 280(4.3) | 102(1.6) | - | 6519(100) | 6,519 | 2809 |
| GLAD | - | 11786(100) | - | 11786(100) | 49(0.4) | - | 11,786 | 11786 |
| MCS - Combined | 3139(13.7) | 206(0.9) | 1125(4.9) | 206(0.9) | 560(2.4) | - | 22,988 | 12427 |
| MCS Members | 1560(15.6) | 138(1.4) | 611(6.1) | 138(1.4) | 326(3.3) | - | 9,980 | |
| MCS Parents | 1579(12.1) | 68(0.5) | 514(4.0) | 68(0.5) | 234(1.8) | - | 13,008 | |
| NCDS58 | 1319(8.1) | 66(0.4) | 362(2.2) | 66(0.4) | 198(1.2) | - | 16,383 | 6963 |
| NextSteps | 1247(15.4) | 112(1.4) | 413(5.1) | 112(1.4) | 208(2.6) | - | 8,084 | 4224 |
| NICOLA | 293(30.4) | - | 56(5.8) | 636(66.0) | - | 963(100) | 963 | 963 |
| TWINSUK | 1196(16.4) | 69(0.9) | 976(13.4) | 69(0.9) | 881(12.1) | - | 7,292 | 3294 |
| UKHLS | 1941(3.9) | 5592(11.2) | 475(1.0) | 5592(11.2) | 372(0.7) | - | 49,736 | 6217 |
| Total | 15174(9.1) | 28378(17.0) | 4970(3.0) | 29099(17.4) | 3053(1.8) | 7482(4.5) | 167,302 | 72290 |

***Table S4 - Cross tabulated self-reported infection, self-reported positive test, and EPR linked positive test.***

| | | Self-reported Positive Test | | | |
|---|---|---|---|---|---|
| | | No | Yes | Missing | Total |
| Self-reported Infection | No | 122,851 | 395 | 504 | 123,750 |
| | Yes | 10,382 | 4,575 | 217 | 15,174 |
| | Missing | - | - | 28,378 | 28,378 |
| | Total | 133,233 | 4,970 | 29,099 | 167,302 |
| | | Linked Positive Test | | | |
| | | No | Yes | Missing | Total |
| Self-reported Infection | No | 115,979 | 1,397 | 6,374 | 123,750 |
| | Yes | 12,615 | 1,468 | 1,091 | 15,174 |
| | Missing | 28,173 | 188 | 17 | 28,378 |
| | Total | 156,767 | 3,053 | 7,482 | 167,302 |
| | | Self-reported Positive Test | | | |
| | | No | Yes | Missing | Total |
| Linked Positive Test | No | 125,242 | 3,352 | 28,173 | 156,767 |
| | Yes | 1,583 | 1,282 | 188 | 3,053 |
| | Missing | 6,408 | 336 | 738 | 7,482 |
| | Total | 133,233 | 4,970 | 29,099 | 167,302 |

***Table S5 – Self-reported infection, self-reported positive test, and EPR linked positive test rates by time-tranche (Apr-Jun2020, Jul-Oct2020, and Nov-Mar2021) and reduced time-tranche (Apr-Oct2020/Nov-Mar2021).***

| Time-Tranche | Self-Reported Infection | | Self-Reported Positive Test | | Linked Positive Test | |
|---|---|---|---|---|---|---|
| | Yes | Total | Yes | Total | Yes | Total |
| Apr20-Jun20 | 4201(9.1) | 46,180 | 582(1.3) | 46,180 | 190(0.3) | 67,070 |
| Jul20-Oct20 | 4263(9.8) | 43,307 | 1241(2.9) | 43,222 | 1210(2.8) | 43,609 |
| Nov20-Mar21 | 6417(13.3) | 48,290 | 3091(6.4) | 48,290 | 1653(3.4) | 48,957 |
| Total | 14881(10.8) | 137,777 | 4914(3.6) | 137,692 | 3053(1.9) | 159,820 |
| Total (inc NICOLA) | 15174(10.9) | 138924 | 4970(3.6) | 138,203 | - | - |
| Reduced Time-tranche | Yes | Total | Yes | Total | Yes | Total |
| Apr20-Oct20 | 8464(9.5) | 89,487 | 1823(2.0) | 89,402 | 1400(1.3) | 110,679 |
| Nov20-Mar21 | 6417(13.3) | 48,290 | 3091(6.4) | 48,290 | 1653(3.4) | 48,957 |
| Total | 14881(10.8) | 137,777 | 4914(3.6) | 137,692 | 3053(1.9) | 159,820 |
| Total (inc NICOLA) | 15174(10.9) | 138924 | 4970(3.6) | 138,203 | - | - |

***Table S5*** **– Self-reported and linked infection rates for Essential occupation by time-tranche, and full study period**

| Self-reported Infection/Time-tranche | Time-tranche 1 March 2020 - October 2020 | | | | Time Tranche-2 Nov 2020 - March 2021 | | | | Full period March 2020 - March 2021 (inc NICOLA) | | | |
|---|---|---|---|---|---|---|---|---|---|---|---|---|
| | Self-reported | | Linked Test Result | | Self-reported | | Linked Test Result | | Self-reported | | Linked Test Result | |
| Age at March 2020 | %Yes | Total | %Yes | Total | %Yes | Total | %Yes | Total | %Yes | Total | %Yes | Total |
| 18-34 | 12.1 | 23,764 | 1.4 | 30,268 | 20.2 | 15,146 | 3.9 | 15,836 | 15.3 | 38,910 | 2.3 | 46,104 |
| 35-44 | 8.2 | 8,974 | 2.1 | 12,963 | 6.5 | 3,458 | 3.7 | 3,753 | 7.7 | 12,432 | 2.5 | 16,716 |
| 45-54 | 9.1 | 20,568 | 1.2 | 25,601 | 11 | 10,576 | 3.3 | 10,750 | 9.8 | 31,144 | 1.8 | 36,351 |
| 55-66 | 7.6 | 27,710 | 1 | 33,340 | 8.2 | 14,085 | 2.6 | 13,561 | 7.8 | 41,795 | 1.5 | 46,901 |
| Total | 9.4 | 81,016 | 1.3 | 102,172 | 13 | 43,265 | 3.3 | 43,900 | 10.6 | 124,281 | 1.9 | 146,072 |
| Sex | %Yes | Total | %Yes | Total | %Yes | Total | %Yes | Total | %Yes | Total | %Yes | Total |
| Female | 9.9 | 57,600 | 1.6 | 72,724 | 13.7 | 29,583 | 3.8 | 29,807 | 11.2 | 87,183 | 2.2 | 102,531 |
| Male | 8.5 | 30,004 | 0.6 | 36,049 | 12.6 | 18,702 | 2.7 | 19,143 | 10.1 | 48,706 | 1.4 | 55,192 |
| Total | 9.4 | 87,604 | 1.3 | 108,773 | 13.3 | 48,285 | 3.4 | 48,950 | 10.8 | 135,889 | 1.9 | 157,723 |
| Ethnicity | %Yes | Total | %Yes | Total | %Yes | Total | %Yes | Total | %Yes | Total | %Yes | Total |
| White | 9.3 | 77,604 | 1.2 | 94,892 | 13.1 | 42,400 | 3.3 | 42,729 | 10.6 | 120,004 | 1.9 | 137,621 |
| Other | 9.6 | 6,990 | 2.2 | 10,791 | 16.3 | 3,421 | 4.5 | 3,706 | 11.8 | 10,411 | 2.8 | 14,497 |
| Total | 9.3 | 84,594 | 1.3 | 105,683 | 13.3 | 45,821 | 3.4 | 46,435 | 10.7 | 130,415 | 1.9 | 152,118 |
| Housing Location | %Yes | Total | %Yes | Total | %Yes | Total | %Yes | Total | %Yes | Total | %Yes | Total |
| Urban | 5 | 20731 | >0.10 | 23274 | 5 | 12489 | <2.26 | 14144 | 5 | 33220 | 0.9 | 37418 |
| Rural | 3.7 | 6960 | <0.15 | 7657 | 2.7 | 4202 | >0.90 | 4643 | 3.3 | 11162 | 0.4 | 12300 |
| Total | 4.7 | 27691 | 0.1 | 30931 | 4.4 | 16691 | 1.9 | 18787 | 4.6 | 44382 | 0.8 | 49718 |
| Housing Composition | %Yes | Total | %Yes | Total | %Yes | Total | %Yes | Total | %Yes | Total | %Yes | Total |
| Partner+Kids | 9.6 | 24797 | 1.9 | 23154 | 11.4 | 12707 | 3.5 | 12167 | 10.2 | 37504 | 2.5 | 35321 |
| Partner Only | 9 | 34724 | 0.8 | 32993 | 12.8 | 19110 | 3.2 | 18474 | 10.3 | 53834 | 1.7 | 51467 |
| Single parent | 11.3 | 4422 | 2 | 4197 | 14.5 | 2370 | 4.5 | 2299 | 12.4 | 6792 | 2.9 | 6496 |
| Other | 8.7 | 14104 | 1.7 | 14203 | 17.6 | 8189 | 3.5 | 8283 | 12 | 22293 | 2.4 | 22486 |
| Alone | 10.1 | 6149 | 0.7 | 5528 | 7.5 | 3523 | 2.5 | 3280 | 9.1 | 9672 | 1.4 | 8808 |
| Total | 9.3 | 84196 | 1.3 | 80075 | 12.9 | 45899 | 3.4 | 44503 | 10.6 | 130095 | 2.1 | 124578 |
| Vaccinations | %Yes | Total | %Yes | Total | %Yes | Total | %Yes | Total | %Yes | Total | %Yes | Total |
| None | >9.0 | 84744 | >1.5 | 88425 | <14 | 30594 | <3.0 | 32863 | 10.4 | 115338 | 1.9 | 121288 |
| One or more | <50 | 22 | <50 | 22 | >14 | 15915 | >4.1 | 16094 | 13.1 | 15937 | 4.2 | 16116 |
| Total | 9.3 | 84766 | 1.5 | 88447 | 13.2 | 46509 | 3.4 | 48957 | 10.7 | 131275 | 2.2 | 137404 |

*Table S6 – Stage 2 combined results of Individual Patient Data meta-analysis, including heterogeneity statistics*

| | | | Self-Reported | | | | | Linked Positive | | | | |
|---|---|---|---|---|---|---|---|---|---|---|---|---|
| Analysis Comparison | Model | Time_Tranche | RR(95%.C.I.) | Q(p-val) | Tau2 | I2 | H2 | RR(95%.C.I.) | Q(p-val) | Tau2 | I2 | H2 |
| Employed?(Unemp v Emp) | Unadj | Overall | 0.89(0.80,1.00) | 0.000 | 0.03 | 76.9 | 4.32 | 0.82(0.69,0.97) | 0.143 | 0.03 | 40.1 | 1.67 |
| Employed?(Unemp v Emp) | Unadj | Apr-Oct20 | 0.86(0.80,0.92) | 0.013 | 0.00 | 0.0 | 1.00 | 0.97(0.88,1.06) | 0.136 | 0.00 | 0.0 | 1.00 |
| Employed?(Unemp v Emp) | Unadj | Nov-Mar21 | 0.91(0.81,1.02) | 0.001 | 0.02 | 58.9 | 2.43 | 0.76(0.66,0.87) | 0.957 | 0.00 | 0.0 | 1.00 |
| Employed?(Unemp v Emp) | Adj Mod1 | Overall | 0.89(0.79,1.00) | 0.000 | 0.03 | 78.3 | 4.62 | 0.82(0.72,0.92) | 0.831 | 0.00 | 0.0 | 1.00 |
| Employed?(Unemp v Emp) | Adj Mod1 | Apr-Oct20 | 0.85(0.79,0.91) | 0.012 | 0.00 | 0.0 | 1.00 | 0.78(0.61,1.01) | 0.109 | 0.05 | 43.8 | 1.78 |
| Employed?(Unemp v Emp) | Adj Mod1 | Nov-Mar21 | 0.91(0.81,1.02) | 0.001 | 0.02 | 57.7 | 2.37 | 0.75(0.65,0.86) | 0.881 | 0.00 | 0.0 | 1.00 |
| Employed?(Unemp v Emp) | Adj Mod2 | Overall | 0.90(0.81,1.00) | 0.000 | 0.02 | 69.7 | 3.30 | 0.81(0.71,0.91) | 0.696 | 0.00 | 0.0 | 1.00 |
| Employed?(Unemp v Emp) | Adj Mod2 | Apr-Oct20 | 0.85(0.79,0.92) | 0.016 | 0.00 | 0.0 | 1.00 | 0.77(0.60,0.98) | 0.126 | 0.05 | 41.7 | 1.71 |
| Employed?(Unemp v Emp) | Adj Mod2 | Nov-Mar21 | 0.92(0.80,1.05) | 0.000 | 0.03 | 68.7 | 3.20 | 0.76(0.66,0.87) | 0.929 | 0.00 | 0.0 | 1.00 |
| Employed?(Retired v Emp) | Unadj | Overall | 0.86(0.66,1.11) | 0.000 | 0.10 | 85.4 | 6.85 | 0.55(0.42,0.73) | 0.635 | 0.00 | 0.0 | 1.00 |
| Employed?(Retired v Emp) | Unadj | Apr-Oct20 | 0.87(0.62,1.22) | 0.000 | 0.14 | 87.3 | 7.86 | 0.63(0.29,1.36) | 0.008 | 0.38 | 71.0 | 3.45 |
| Employed?(Retired v Emp) | Unadj | Nov-Mar21 | 1.03(0.52,2.04) | 0.000 | 0.68 | 97.1 | 34.68 | 0.59(0.40,0.87) | 0.453 | 0.00 | 0.0 | 1.00 |
| Employed?(Retired v Emp) | Adj Mod1 | Overall | 0.88(0.72,1.08) | 0.000 | 0.06 | 75.9 | 4.15 | 0.57(0.43,0.75) | 0.715 | 0.00 | 0.0 | 1.00 |
| Employed?(Retired v Emp) | Adj Mod1 | Apr-Oct20 | 0.94(0.69,1.28) | 0.000 | 0.11 | 82.9 | 5.86 | 0.66(0.30,1.46) | 0.011 | 0.40 | 70.9 | 3.44 |
| Employed?(Retired v Emp) | Adj Mod1 | Nov-Mar21 | 1.11(0.60,2.05) | 0.000 | 0.55 | 96.2 | 26.57 | 0.60(0.40,0.88) | 0.682 | 0.00 | 0.0 | 1.00 |
| Employed?(Retired v Emp) | Adj Mod2 | Overall | 0.88(0.73,1.04) | 0.001 | 0.04 | 65.7 | 2.92 | 0.58(0.44,0.77) | 0.772 | 0.00 | 0.0 | 1.00 |
| Employed?(Retired v Emp) | Adj Mod2 | Apr-Oct20 | 0.94(0.66,1.34) | 0.000 | 0.12 | 84.7 | 6.55 | 0.86(0.45,1.64) | 0.136 | 0.19 | 50.3 | 2.01 |
| Employed?(Retired v Emp) | Adj Mod2 | Nov-Mar21 | 1.34(0.57,3.18) | 0.000 | 0.91 | 95.3 | 21.35 | 0.62(0.42,0.92) | 0.657 | 0.00 | 0.0 | 1.00 |
| Emp Status(Part-time v FT) | Unadj | Overall | 0.95(0.90,1.00) | 0.661 | 0.00 | 0.0 | 1.00 | 0.86(0.69,1.06) | 0.056 | 0.05 | 47.6 | 1.91 |
| Emp Status(Part-time v FT) | Unadj | Apr-Oct20 | 0.87(0.79,0.95) | 0.317 | 0.00 | 20.5 | 1.26 | 0.63(0.47,0.85) | 0.467 | 0.02 | 10.3 | 1.11 |
| Emp Status(Part-time v FT) | Unadj | Nov-Mar21 | 0.98(0.86,1.11) | 0.023 | 0.02 | 52.9 | 2.12 | 0.95(0.78,1.17) | 0.428 | 0.01 | 15.0 | 1.18 |
| Emp Status(Part-time v FT) | Adj Mod1 | Overall | 0.94(0.89,0.99) | 0.480 | 0.00 | 0.0 | 1.00 | 0.82(0.65,1.04) | 0.020 | 0.08 | 55.8 | 2.26 |
| Emp Status(Part-time v FT) | Adj Mod1 | Apr-Oct20 | 0.86(0.78,0.94) | 0.254 | 0.00 | 20.9 | 1.26 | 0.59(0.43,0.81) | 0.549 | 0.03 | 14.0 | 1.16 |
| Emp Status(Part-time v FT) | Adj Mod1 | Nov-Mar21 | 0.96(0.85,1.09) | 0.033 | 0.02 | 50.8 | 2.03 | 0.92(0.73,1.14) | 0.261 | 0.03 | 27.8 | 1.38 |
| Emp Status(Part-time v FT) | Adj Mod2 | Overall | 0.96(0.90,1.02) | 0.284 | 0.00 | 16.3 | 1.19 | 0.89(0.71,1.11) | 0.141 | 0.05 | 41.5 | 1.71 |
| Emp Status(Part-time v FT) | Adj Mod2 | Apr-Oct20 | 0.87(0.79,0.95) | 0.190 | 0.00 | 23.8 | 1.31 | 0.68(0.50,0.93) | 0.722 | 0.00 | 0.0 | 1.00 |
| Emp Status(Part-time v FT) | Adj Mod2 | Nov-Mar21 | 0.97(0.86,1.09) | 0.053 | 0.01 | 47.2 | 1.89 | 0.90(0.72,1.13) | 0.347 | 0.03 | 23.2 | 1.30 |
| Emp Status(Emp but not wking v FT) | Unadj | Overall | 0.99(0.89,1.11) | 0.000 | 0.03 | 71.7 | 3.54 | 1.05(0.81,1.35) | 0.001 | 0.10 | 60.4 | 2.53 |
| Emp Status(Emp but not wking v FT) | Unadj | Apr-Oct20 | 0.92(0.75,1.13) | 0.000 | 0.09 | 85.6 | 6.92 | 0.93(0.66,1.30) | 0.055 | 0.03 | 10.6 | 1.12 |

| | | | | | | | | | | | | |
|---|---|---|---|---|---|---|---|---|---|---|---|---|
| Emp Status(Emp but not wking v FT) | Unadj | Nov-Mar21 | 1.06(0.91,1.23) | 0.001 | 0.04 | 71.3 | 3.48 | 1.08(0.81,1.43) | 0.008 | 0.10 | 56.9 | 2.32 |
| Emp Status(Emp but not wking v FT) | Adj Mod1 | Overall | 0.99(0.88,1.11) | 0.000 | 0.03 | 72.4 | 3.62 | 1.03(0.79,1.33) | 0.001 | 0.10 | 59.9 | 2.49 |
| Emp Status(Emp but not wking v FT) | Adj Mod1 | Apr-Oct20 | 0.92(0.75,1.13) | 0.000 | 0.10 | 85.6 | 6.93 | 0.90(0.68,1.20) | 0.071 | 0.00 | 0.0 | 1.00 |
| Emp Status(Emp but not wking v FT) | Adj Mod1 | Nov-Mar21 | 1.05(0.90,1.22) | 0.001 | 0.04 | 70.7 | 3.41 | 1.05(0.79,1.40) | 0.010 | 0.09 | 55.7 | 2.26 |
| Emp Status(Emp but not wking v FT) | Adj Mod2 | Overall | 0.98(0.88,1.09) | 0.000 | 0.02 | 68.5 | 3.18 | 1.02(0.80,1.29) | 0.006 | 0.07 | 52.4 | 2.10 |
| Emp Status(Emp but not wking v FT) | Adj Mod2 | Apr-Oct20 | 0.92(0.74,1.15) | 0.000 | 0.10 | 86.2 | 7.27 | 0.85(0.63,1.15) | 0.288 | 0.00 | 0.0 | 1.00 |
| Emp Status(Emp but not wking v FT) | Adj Mod2 | Nov-Mar21 | 1.03(0.88,1.21) | 0.002 | 0.04 | 70.6 | 3.40 | 1.05(0.81,1.38) | 0.029 | 0.08 | 50.1 | 2.00 |
| Emp Status(Unempl v FT) | Unadj | Overall | 0.85(0.73,1.00) | 0.000 | 0.06 | 84.5 | 6.46 | 0.75(0.61,0.92) | 0.032 | 0.05 | 48.9 | 1.96 |
| Emp Status(Unempl v FT) | Unadj | Apr-Oct20 | 0.75(0.63,0.90) | 0.000 | 0.07 | 75.6 | 4.09 | 0.53(0.30,0.94) | 0.000 | 0.45 | 68.1 | 3.13 |
| Emp Status(Unempl v FT) | Unadj | Nov-Mar21 | 0.91(0.81,1.03) | 0.020 | 0.02 | 55.2 | 2.23 | 0.74(0.63,0.87) | 0.776 | 0.00 | 0.0 | 1.00 |
| Emp Status(Unempl v FT) | Adj Mod1 | Overall | 0.85(0.72,1.01) | 0.000 | 0.07 | 85.9 | 7.11 | 0.73(0.62,0.86) | 0.434 | 0.01 | 17.5 | 1.21 |
| Emp Status(Unempl v FT) | Adj Mod1 | Apr-Oct20 | 0.74(0.61,0.90) | 0.000 | 0.07 | 76.1 | 4.19 | 0.53(0.31,0.90) | 0.000 | 0.35 | 60.6 | 2.54 |
| Emp Status(Unempl v FT) | Adj Mod1 | Nov-Mar21 | 0.91(0.80,1.02) | 0.031 | 0.02 | 52.5 | 2.11 | 0.71(0.61,0.84) | 0.693 | 0.00 | 0.0 | 1.00 |
| Emp Status(Unempl v FT) | Adj Mod2 | Overall | 0.84(0.72,0.97) | 0.000 | 0.05 | 81.4 | 5.39 | 0.72(0.63,0.83) | 0.599 | 0.00 | 0.0 | 1.00 |
| Emp Status(Unempl v FT) | Adj Mod2 | Apr-Oct20 | 0.76(0.62,0.93) | 0.001 | 0.07 | 75.6 | 4.10 | 0.56(0.37,0.85) | 0.188 | 0.11 | 30.5 | 1.44 |
| Emp Status(Unempl v FT) | Adj Mod2 | Nov-Mar21 | 0.91(0.79,1.06) | 0.002 | 0.03 | 67.2 | 3.05 | 0.73(0.62,0.87) | 0.715 | 0.00 | 0.0 | 1.00 |
| Emp Status(Retired v FT) | Unadj | Overall | 0.80(0.62,1.02) | 0.000 | 0.10 | 81.3 | 5.35 | 0.54(0.34,0.87) | 0.008 | 0.18 | 73.4 | 3.75 |
| Emp Status(Retired v FT) | Unadj | Apr-Oct20 | 0.75(0.56,1.01) | 0.000 | 0.14 | 82.1 | 5.59 | 0.87(0.64,1.20) | 0.109 | 0.05 | 58.0 | 2.38 |
| Emp Status(Retired v FT) | Unadj | Nov-Mar21 | 0.98(0.56,1.69) | 0.000 | 0.57 | 96.3 | 27.16 | 0.64(0.47,0.86) | 0.521 | 0.00 | 0.0 | 1.00 |
| Emp Status(Retired v FT) | Adj Mod1 | Overall | 0.80(0.66,0.98) | 0.001 | 0.06 | 69.8 | 3.31 | 0.58(0.38,0.90) | 0.020 | 0.13 | 67.6 | 3.08 |
| Emp Status(Retired v FT) | Adj Mod1 | Apr-Oct20 | 0.79(0.61,1.03) | 0.001 | 0.10 | 75.1 | 4.01 | 0.91(0.66,1.27) | 0.158 | 0.07 | 56.5 | 2.30 |
| Emp Status(Retired v FT) | Adj Mod1 | Nov-Mar21 | 1.08(0.66,1.79) | 0.000 | 0.46 | 95.2 | 20.91 | 0.69(0.48,1.01) | 0.358 | 0.03 | 17.2 | 1.21 |
| Emp Status(Retired v FT) | Adj Mod2 | Overall | 0.81(0.68,0.96) | 0.004 | 0.04 | 59.5 | 2.47 | 0.71(0.54,0.94) | 0.384 | 0.03 | 30.4 | 1.44 |
| Emp Status(Retired v FT) | Adj Mod2 | Apr-Oct20 | 0.82(0.64,1.06) | 0.001 | 0.09 | 72.6 | 3.65 | 1.02(0.86,1.19) | 0.603 | 0.00 | 5.8 | 1.06 |
| Emp Status(Retired v FT) | Adj Mod2 | Nov-Mar21 | 1.21(0.63,2.30) | 0.000 | 0.67 | 93.2 | 14.63 | 0.73(0.48,1.11) | 0.287 | 0.05 | 26.6 | 1.36 |
| Home wking(Some hm wrk vs all) | Unadj | Overall | 1.06(0.97,1.17) | 0.188 | 0.01 | 38.0 | 1.61 | 1.40(1.16,1.70) | 0.632 | 0.00 | 0.0 | 1.00 |
| Home wking(Some hm wrk vs all) | Unadj | Apr-Oct20 | 1.04(0.92,1.17) | 0.170 | 0.01 | 40.8 | 1.69 | 1.84(0.96,3.52) | 0.021 | 0.45 | 63.1 | 2.71 |
| Home wking(Some hm wrk vs all) | Unadj | Nov-Mar21 | 1.08(0.95,1.23) | 0.022 | 0.02 | 52.3 | 2.10 | 1.35(1.11,1.66) | 0.477 | 0.00 | 0.0 | 1.00 |
| Home wking(Some hm wrk vs all) | Adj Mod1 | Overall | 1.06(0.97,1.17) | 0.233 | 0.01 | 35.2 | 1.54 | 1.39(1.14,1.69) | 0.647 | 0.00 | 0.0 | 1.00 |
| Home wking(Some hm wrk vs all) | Adj Mod1 | Apr-Oct20 | 1.04(0.92,1.17) | 0.206 | 0.01 | 39.2 | 1.64 | 1.96(0.92,4.16) | 0.013 | 0.63 | 62.3 | 2.65 |
| Home wking(Some hm wrk vs all) | Adj Mod1 | Nov-Mar21 | 1.08(0.95,1.23) | 0.032 | 0.02 | 50.6 | 2.02 | 1.33(1.08,1.63) | 0.499 | 0.00 | 0.0 | 1.00 |
| Home wking(Some hm wrk vs all) | Adj Mod2 | Overall | 1.08(1.01,1.16) | 0.574 | 0.00 | 2.8 | 1.03 | 1.40(1.15,1.71) | 0.591 | 0.00 | 0.0 | 1.00 |
| Home wking(Some hm wrk vs all) | Adj Mod2 | Apr-Oct20 | 1.04(0.93,1.17) | 0.366 | 0.01 | 24.1 | 1.32 | 1.76(0.90,3.44) | 0.010 | 0.57 | 66.1 | 2.95 |

| | | | | | | | | | | | |
|---|---|---|---|---|---|---|---|---|---|---|---|
| Home wking(Some hm wrk vs all) | Adj Mod2 | Nov-Mar21 | 1.11(0.99,1.24) | 0.173 | 0.01 | 32.5 | 1.48 | 1.33(1.08,1.64) | 0.490 | 0.00 | 0.0 | 1.00 |
| Home wking(Non hm wrk v all) | Unadj | Overall | 1.08(0.98,1.19) | 0.001 | 0.01 | 64.6 | 2.82 | 1.79(1.44,2.23) | 0.038 | 0.06 | 51.1 | 2.05 |
| Home wking(Non hm wrk v all) | Unadj | Apr-Oct20 | 1.05(0.93,1.18) | 0.000 | 0.02 | 68.0 | 3.12 | 2.98(1.80,4.92) | 0.003 | 0.30 | 67.5 | 3.07 |
| Home wking(Non hm wrk v all) | Unadj | Nov-Mar21 | 1.09(0.94,1.27) | 0.000 | 0.04 | 79.4 | 4.84 | 1.63(1.33,2.00) | 0.161 | 0.03 | 34.3 | 1.52 |
| Home wking(Non hm wrk v all) | Adj Mod1 | Overall | 1.08(0.98,1.19) | 0.001 | 0.02 | 66.9 | 3.02 | 1.78(1.42,2.22) | 0.015 | 0.06 | 52.2 | 2.09 |
| Home wking(Non hm wrk v all) | Adj Mod1 | Apr-Oct20 | 1.04(0.93,1.18) | 0.000 | 0.02 | 66.7 | 3.00 | 3.13(1.81,5.42) | 0.001 | 0.36 | 68.8 | 3.20 |
| Home wking(Non hm wrk v all) | Adj Mod1 | Nov-Mar21 | 1.11(0.95,1.28) | 0.000 | 0.04 | 77.3 | 4.40 | 1.64(1.37,1.96) | 0.196 | 0.01 | 17.6 | 1.21 |
| Home wking(Non hm wrk v all) | Adj Mod2 | Overall | 1.03(0.91,1.16) | 0.000 | 0.03 | 76.8 | 4.31 | 1.77(1.40,2.25) | 0.025 | 0.07 | 55.3 | 2.24 |
| Home wking(Non hm wrk v all) | Adj Mod2 | Apr-Oct20 | 1.04(0.91,1.19) | 0.000 | 0.03 | 68.5 | 3.17 | 3.23(1.76,5.93) | 0.000 | 0.49 | 73.8 | 3.82 |
| Home wking(Non hm wrk v all) | Adj Mod2 | Nov-Mar21 | 1.16(1.01,1.32) | 0.001 | 0.03 | 68.7 | 3.19 | 1.62(1.35,1.96) | 0.240 | 0.02 | 21.1 | 1.27 |
| Keyworker (Yes v No) | Unadj | Overall | 1.24(1.17,1.32) | 0.040 | 0.00 | 39.6 | 1.66 | 1.87(1.55,2.26) | 0.000 | 0.07 | 66.2 | 2.96 |
| Keyworker (Yes v No) | Unadj | Apr-Oct20 | 1.21(1.14,1.28) | 0.573 | 0.00 | 0.0 | 1.00 | 2.30(1.58,3.36) | 0.000 | 0.27 | 76.0 | 4.16 |
| Keyworker (Yes v No) | Unadj | Nov-Mar21 | 1.30(1.16,1.46) | 0.000 | 0.02 | 67.1 | 3.04 | 1.57(1.36,1.81) | 0.308 | 0.01 | 13.6 | 1.16 |
| Keyworker (Yes v No) | Adj Mod1 | Overall | 1.24(1.17,1.32) | 0.045 | 0.00 | 37.3 | 1.59 | 1.85(1.56,2.20) | 0.000 | 0.05 | 58.0 | 2.38 |
| Keyworker (Yes v No) | Adj Mod1 | Apr-Oct20 | 1.19(1.12,1.27) | 0.469 | 0.00 | 2.3 | 1.02 | 2.32(1.55,3.49) | 0.000 | 0.33 | 70.8 | 3.43 |
| Keyworker (Yes v No) | Adj Mod1 | Nov-Mar21 | 1.30(1.16,1.46) | 0.000 | 0.02 | 65.5 | 2.90 | 1.64(1.42,1.90) | 0.551 | 0.00 | 0.0 | 1.00 |
| Keyworker (Yes v No) | Adj Mod2 | Overall | 1.24(1.18,1.31) | 0.191 | 0.00 | 17.3 | 1.21 | 1.65(1.45,1.87) | 0.187 | 0.01 | 25.6 | 1.34 |
| Keyworker (Yes v No) | Adj Mod2 | Apr-Oct20 | 1.17(1.10,1.26) | 0.204 | 0.00 | 15.3 | 1.18 | 1.77(1.25,2.51) | 0.011 | 0.17 | 55.7 | 2.26 |
| Keyworker (Yes v No) | Adj Mod2 | Nov-Mar21 | 1.32(1.16,1.49) | 0.000 | 0.03 | 67.9 | 3.12 | 1.61(1.37,1.89) | 0.393 | 0.01 | 8.2 | 1.09 |
| Furlough (Yes v No) | Unadj | Overall | 0.93(0.88,0.99) | 0.482 | 0.00 | 0.0 | 1.00 | 0.83(0.68,1.00) | 0.737 | 0.00 | 0.0 | 1.00 |
| Furlough (Yes v No) | Unadj | Apr-Oct20 | 0.84(0.72,0.98) | 0.001 | 0.04 | 66.5 | 2.99 | 0.76(0.52,1.11) | 0.510 | 0.00 | 0.0 | 1.00 |
| Furlough (Yes v No) | Unadj | Nov-Mar21 | 1.03(0.94,1.12) | 0.479 | 0.00 | 0.0 | 1.00 | 0.83(0.66,1.05) | 0.604 | 0.00 | 1.0 | 1.01 |
| Furlough (Yes v No) | Adj Mod1 | Overall | 0.93(0.88,0.99) | 0.370 | 0.00 | 0.0 | 1.00 | 0.80(0.66,0.98) | 0.849 | 0.00 | 0.0 | 1.00 |
| Furlough (Yes v No) | Adj Mod1 | Apr-Oct20 | 0.84(0.72,0.98) | 0.001 | 0.05 | 67.0 | 3.03 | 0.78(0.53,1.14) | 0.594 | 0.00 | 0.0 | 1.00 |
| Furlough (Yes v No) | Adj Mod1 | Nov-Mar21 | 1.02(0.93,1.11) | 0.561 | 0.00 | 0.0 | 1.00 | 0.80(0.64,1.01) | 0.824 | 0.00 | 0.0 | 1.00 |
| Furlough (Yes v No) | Adj Mod2 | Overall | 0.90(0.85,0.96) | 0.727 | 0.00 | 0.0 | 1.00 | 0.82(0.67,1.01) | 0.835 | 0.00 | 0.0 | 1.00 |
| Furlough (Yes v No) | Adj Mod2 | Apr-Oct20 | 0.84(0.72,0.98) | 0.001 | 0.05 | 66.0 | 2.94 | 0.77(0.53,1.13) | 0.600 | 0.00 | 0.0 | 1.00 |
| Furlough (Yes v No) | Adj Mod2 | Nov-Mar21 | 0.97(0.88,1.05) | 0.707 | 0.00 | 0.0 | 1.00 | 0.82(0.65,1.04) | 0.691 | 0.00 | 0.0 | 1.00 |

### S7. Appendices – Example Forest-plots (Furlough vs Not)

#### *Self-reported – Unadjusted models*

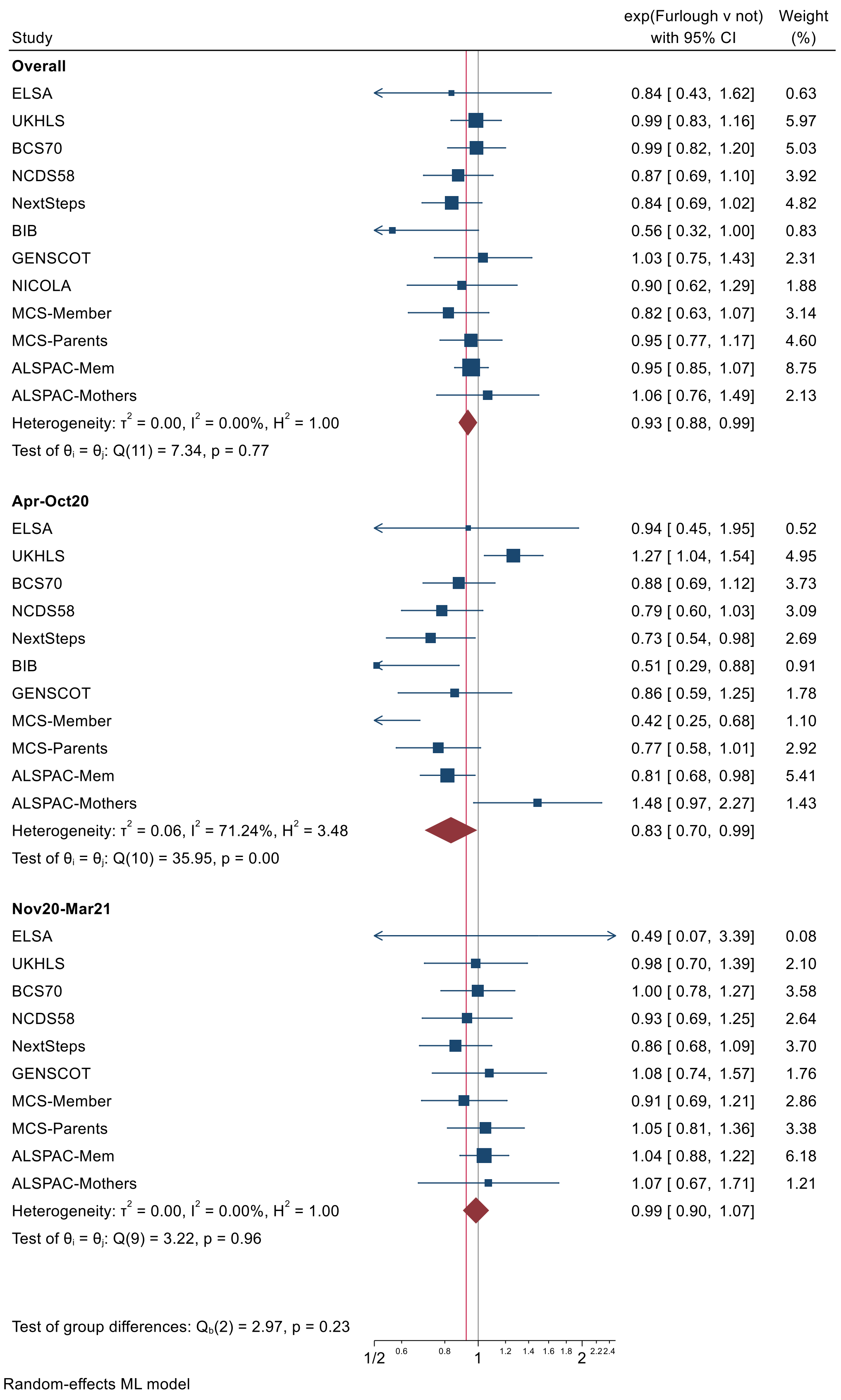

***Self-reported – Adjusted models***

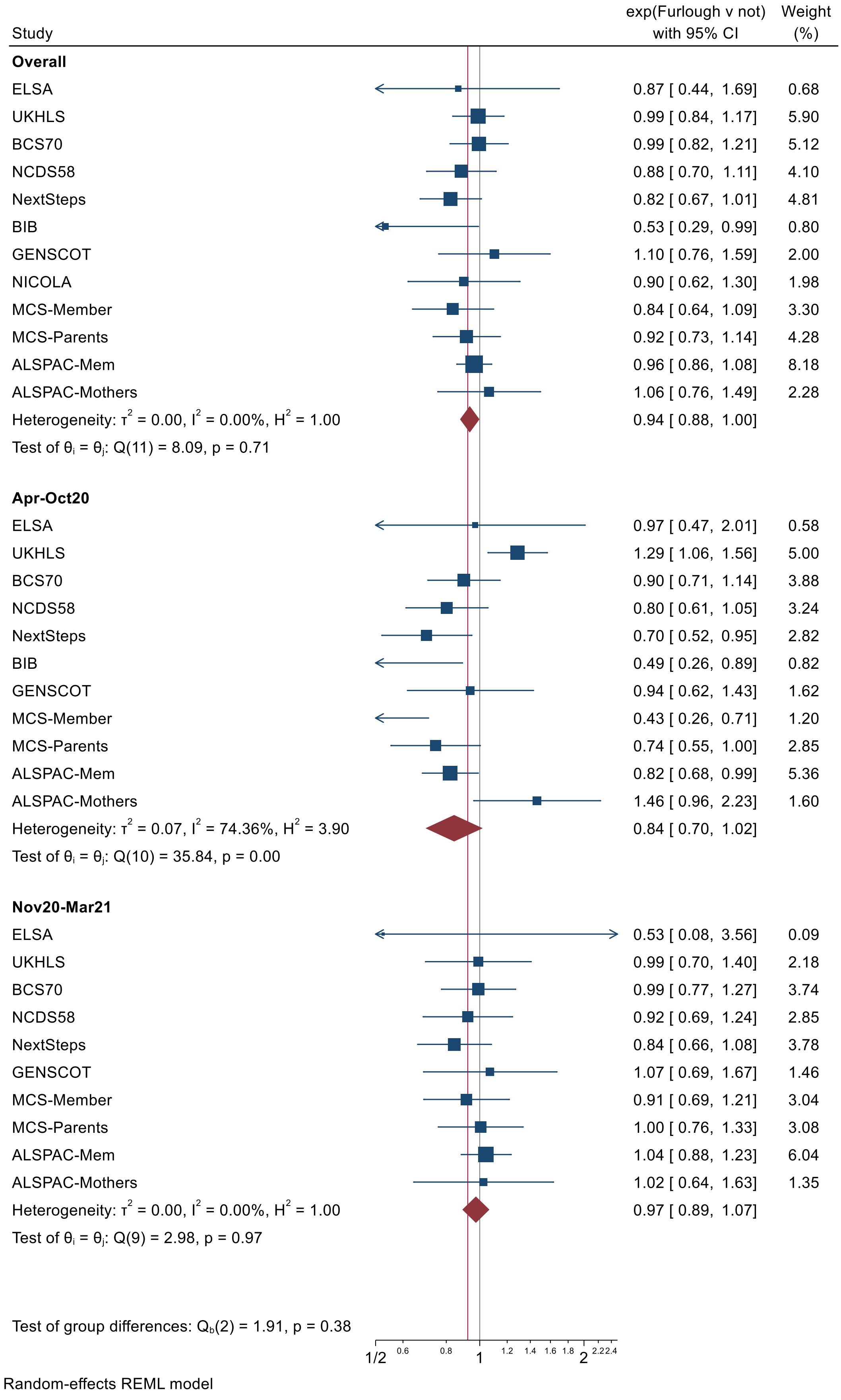

***Linked positive – Unadjusted models***

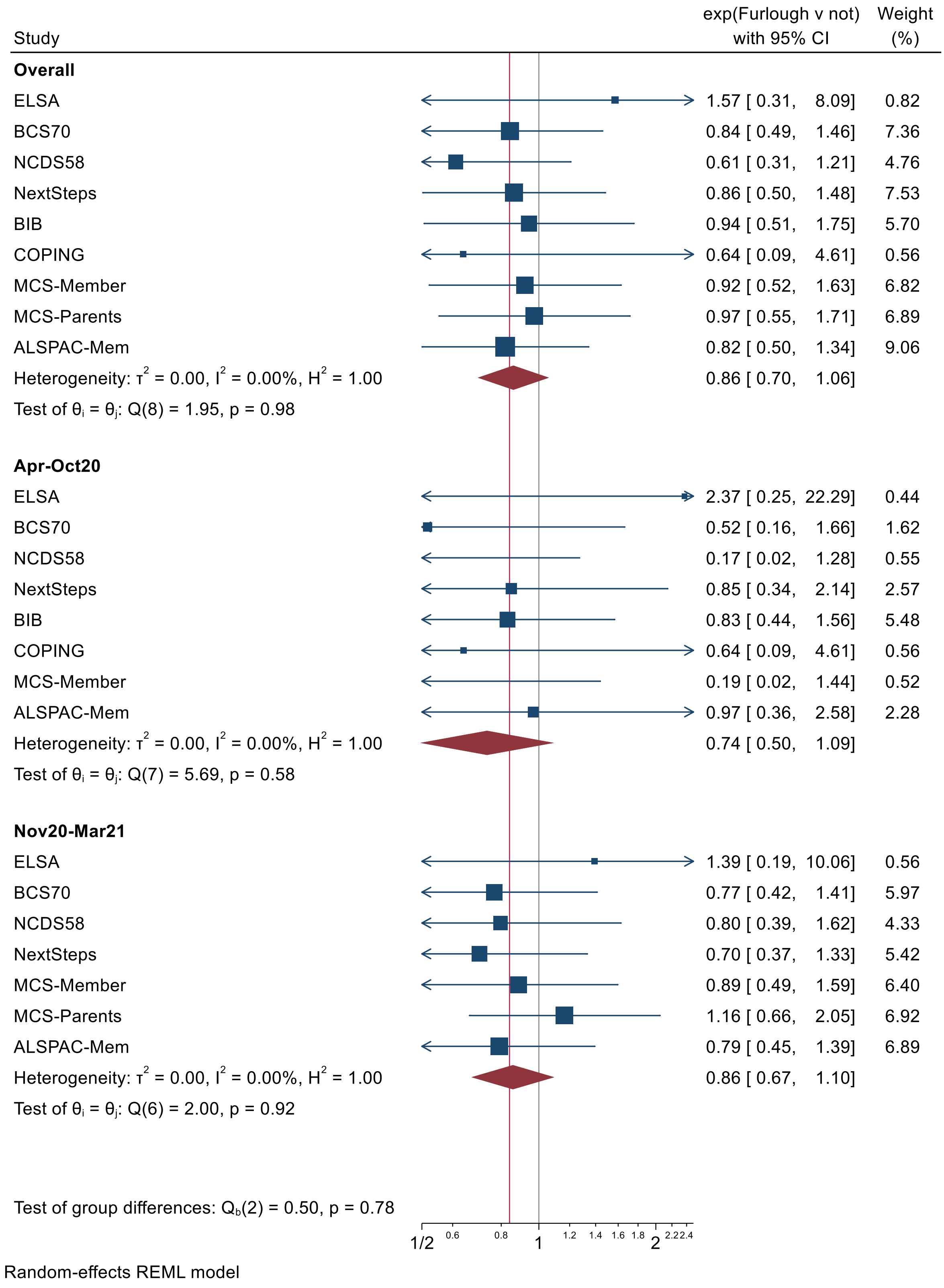

#### *Linked Positive – Adjusted models*

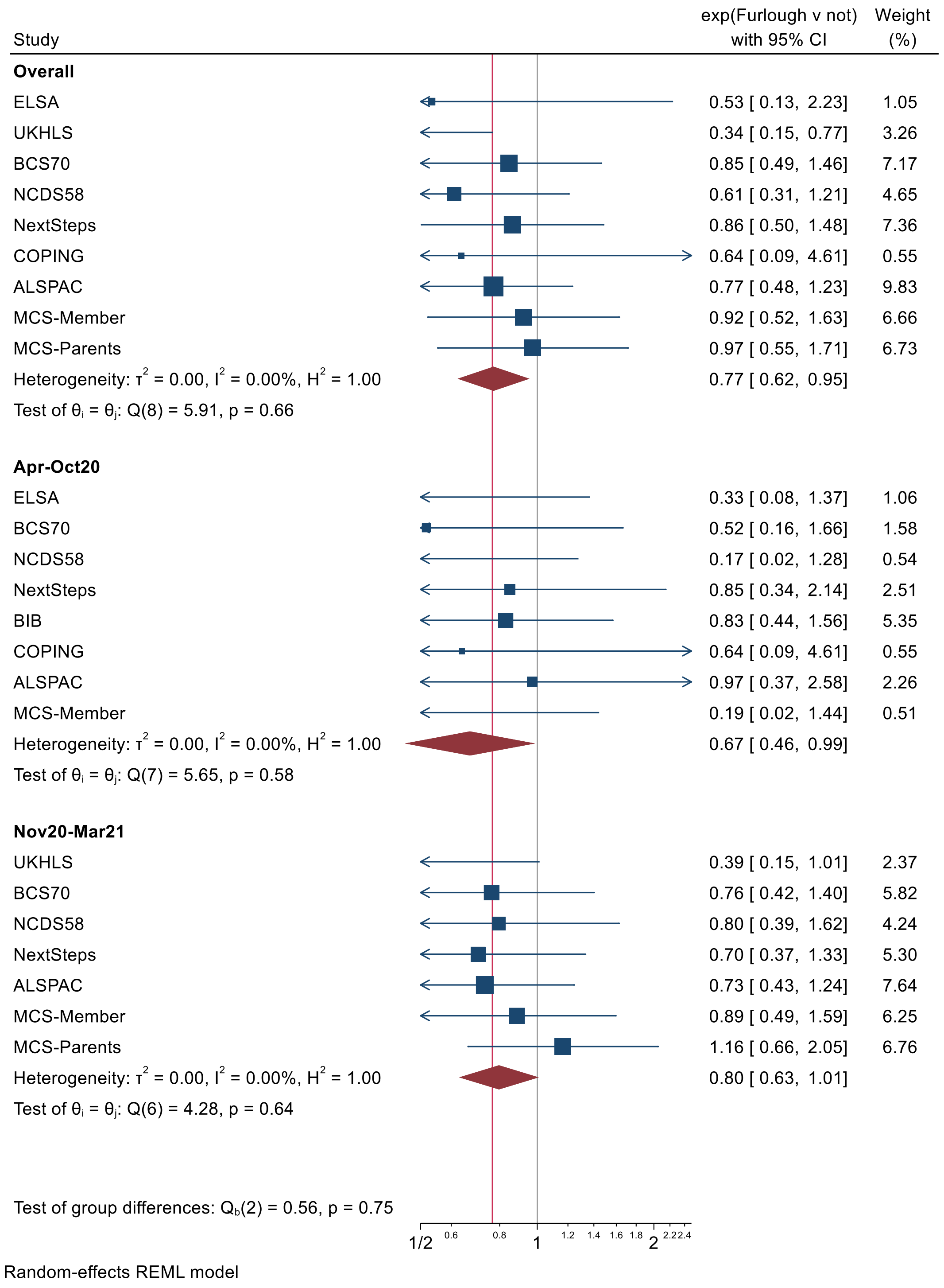

### S8 - Information about Longitudinal Population Studies (LPS) for researchers to include in the Supplementary Materials of their publications

| | | | |
|---|---|---|---|
| **DOC number:** | DOC-ISM-032 | **Version:** | V1.1 |
| **Author(s):** | Katharine Evans, UK LLC Governance & Policy Manager | **Date:** | 27/09/2022 |
| **Authorised by:** | Andy Boyd, UK LLC Director | **Date:** | 08/11/2022 |
| **Date published:** | 09/11/2022 | **Date to review:** | 20/12/2023 |
| **Permission to edit this document must be provided by:** | Director; Senior Research Manager | | |

**Review History**

| Version | Review Date | Reviewed by | Section(s) amended | Authorised by |
|---|---|---|---|---|
| V1.1 | 20/12/2022 | Katharine Evans | Added websites for UKHLS data requests | Andy Boyd, Director |

#### INTRODUCTION

The UK LLC Publication Policy describes the **requirements** for researchers when publishing **papers and similar outputs (such as reports, books and other manuscripts)** based on data accessed in the UK LLC Trusted Research Environment (TRE).

More than 20 LPS provide data to the UK LLC TRE and researchers **must** acknowledge each LPS included in the analyses that underpin their publications – **see the** UK LLC Publication Policy **for the required wording for your Methods and Acknowledgements sections**.

**The information in the tables below has been provided by the LPS**. Copy the information about each LPS that has contributed to your research and include it in the **Supplementary Materials** section of your publication – **you do not have to present the information in table format**.

#### LPS INFORMATION **(LPS listed in alphabetical order)**

| ALSPAC: Avon Longitudinal Study of Parents and Children | |
|---|---|
| **Description of Study Population** (including citations and references if required) | Pregnant women resident in a defined area of the former county of Avon, UK with expected dates of delivery 1st April 1991 to 31st December 1992 were invited to take part in the study [1,2]. The initial number of pregnancies enrolled is 14,541 (14,676 foetuses), resulting in 14,062 live births and 13,988 children who were alive at 1 year of age. Further recruitment took place after the age of 7 years, the total sample size for analyses using any data collected after the age of 7 is therefore 15,454 pregnancies, resulting in 15,589 foetuses. Of these 14,901 were alive at 1 year of age.<br><br>[1]Boyd A, Golding J, Macleod J, Lawlor DA, Fraser A, Henderson J, Molloy L, Ness A, Ring S, Davey Smith G. Cohort Profile: The 'Children of the 90s'; the index offspring of The Avon Longitudinal Study of Parents and Children (ALSPAC). International Journal of Epidemiology 2013; 42: 111-127.<br><br>[2]Fraser A, Macdonald-Wallis C, Tilling K, Boyd A, Golding J, Davey Smith G, Henderson J, Macleod J, Molloy L, Ness A, Ring S, Nelson SM, Lawlor DA. Cohort Profile: The Avon Longitudinal Study of Parents and Children: ALSPAC mothers cohort. International Journal of Epidemiology 2013; 42:97-110. |
| **Acknowledgements** | We are extremely grateful to all the families who took part in this study, the midwives for their help in recruiting them, and |

| | |
|---|---|
| | the whole ALSPAC team, which includes interviewers, computer and laboratory technicians, clerical workers, research scientists, volunteers, managers, receptionists and nurses. |
| **Ethics** | Ethical approval for the study was obtained from the ALSPAC Law and Ethics committee and local research ethics committees (NHS Haydock REC: 10/H1010/70). |
| **Website for Data Requests** | http://www.bristol.ac.uk/alspac/researchers/access/ |

| BCS70: 1970 British Cohort Study | |
|---|---|
| **Description of Study Population** (including citations and references if required) | The 1970 British Cohort Study (BCS70) follows the lives of more than 17,000 people born in England, Scotland and Wales in a single week of 1970. Over the course of cohort members' lives, BCS70 has collected information on health, physical, educational and social development, and economic circumstances, among other factors.<br><br>Since the birth survey in 1970, there have been nine 'sweeps' of all cohort members at ages 5, 10, 16, 26, 30, 34, 38, 42 and most recently at 46 (a biomedical data collection). The Age 51 Sweep is currently in the field (2022).<br><br>Data have been collected from a number of different sources, including the midwife present at birth, parents of the cohort members, head and class teachers, school health service personnel and the cohort members themselves.<br><br>The data have been collected in a variety of ways, including via paper and electronic questionnaires, clinical records, medical examinations, biological samples, physical measurements, tests of ability, educational assessments and diaries. |

| | |
|---|---|
| | The study is conducted by the Centre for Longitudinal Studies. |
| **Acknowledgements** | BCS70 is core-funded by the ESRC. |
| **Ethics** | Ethics approval has been obtained for each follow-up from an NHS Research Ethics Committee (REC) since 2000. In addition, separate REC approval is in place to cover the ongoing activities of the study in between major sweeps of data collection (i.e. Keeping in touch with and tracing cohort members; cleaning, documenting and providing access to the data for research; and linking data from administrative sources to survey data to increase the utility of the data for research). |
| **Website for Data Requests** | https://cls.ucl.ac.uk/cls-studies/bcs70/ |

| **BIB: Born in Bradford** | |
|---|---|
| **Description of Study Population** (including citations and references if required) | Born in Bradford (BiB) is a prospective pregnancy and birth cohort study of the children, mothers and fathers from 13,776 pregnancies from between 2007 and 2011, based in Bradford, UK. The study was established to examine how genetic, nutritional, environmental, behavioural and social factors affect health and development during childhood, and subsequently, adult life in a deprived multi-ethnic population[1]. From 2017 to 2021 a full follow-up of the cohort was conducted[2] to investigate the determinants of children's pre-pubertal health and development, including through understanding parents' health and wellbeing, and to obtain data on exposures in childhood that might influence future health. In 2022, the Age of Wonder study was launched to complete another full follow-up through adolescence into adulthood, focusing on priority areas of physical and mental health, growth, identity, cognition, socioeconomic status and environmental exposures.<br><br>[1]John Wright, Neil Small, Pauline Raynor, Derek Tuffnell, Raj Bhopal, Noel Cameron, Lesley Fairley, Debbie A Lawlor, Roger Parslow, Emily S Petherick, Kate E Pickett, Dagmar |

| | |
|---|---|
| | Waiblinger, Jane West, on behalf of the Born in Bradford Scientific Collaborators Group, Cohort Profile: The Born in Bradford multi-ethnic family cohort study, International Journal of Epidemiology, Volume 42, Issue 4, August 2013, Pages 978–991, https://doi.org/10.1093/ije/dys112<br>[2]Bird, P.K., McEachan, R.R.C., Mon-Williams, M. et al. Growing up in Bradford: protocol for the age 7–11 follow up of the Born in Bradford birth cohort. BMC Public Health 19, 939 (2019). https://doi.org/10.1186/s12889-019-7222-2 |
| **Acknowledgements** | Born in Bradford has received funding from the Wellcome Trust [101597/Z/13/Z and 223601/Z/21/Z]; a joint grant from the UK Medical Research Council (MRC) and UK Economic and Social Science Research Council (ESRC) [MR/N024391/1; a British Heart Foundation Clinical Study grant [CS/16/4/32482]; the National Institute for Health Research under its Applied Research Collaboration Yorkshire and Humber [NIHR200166]; and a Health Foundation COVID-19 Award [2301201]. Born in Bradford is only possible because of the enthusiasm and commitment of the Children and Parents in BiB. We are grateful to all the participants, health professionals, schools and researchers who have made Born in Bradford happen.<br><br>**NMR metabolomics data – additional acknowledgement**<br>Researchers who use the NMR metabolomics data should cite the following data note:<br>https://wellcomeopenresearch.org/articles/5-264<br>The funding acknowledgement should be:<br>Funding for the metabolomics analyses in BiB has been provided by the US National Institutes of Health [R01 DK10324]; the European Research Council (ERC) under the European Union's Seventh Framework Programme [FP7/2007-2013] / ERC grant agreement no 669545; and the UK Medical Research Council [MC_UU_00011/6]. |
| **Ethics** | Study: Born in Bradford Age of Wonder: a co-produced mixed methods longitudinal exploration of health and wellbeing trajectories through adolescence and young adulthood in the multi-cultural city of Bradford, UK.<br>REC: 21/YH/0261<br><br>Study: Born in Bradford's Growing up Family Study<br>REC: 16/YH/0320 |

| | |
|---|---|
| | Study: Born in Bradford: A longitudinal cohort study of babies born in Bradford and their mothers and fathers<br>REC: 07/H1302/112 |
| **Website for Data Requests** | https://www.borninbradford.nhs.uk |

| **ELSA: English Longitudinal Study of Ageing** | |
|---|---|
| **Description of Study Population** (including citations and references if required) | The English Longitudinal Study of Ageing (ELSA) is a unique and rich resource of information on the dynamics of health, social, wellbeing and economic circumstances in the English population aged 50 and older [1].<br><br>The original sample was drawn from households that had previously responded to the Health Survey for England (HSE) between 1998 and 2001. The main fieldwork began in March 2002. The same group of respondents have been interviewed at two-yearly interviews .<br><br>[1]Banks J, Batty GD, Breedvelt JJF, Coughlin K, Crawford R, Marmot M, Nazroo J, Oldfield Z, Steel N, Steptoe A, Wood M, Zaninotto P (2021) English Longitudinal Study of Ageing: Waves 0-9, 1998-2019 |
| **Acknowledgements** | The English Longitudinal Study of Ageing was developed by a team of researchers based at University College London, NatCen Social Research, the Institute for Fiscal Studies, the University of Manchester and the University of East Anglia. The data were collected by NatCen Social Research. The funding is currently provided by the National Institute on Aging (Ref: R01AG017644) and by a consortium of UK government departments: Department for Health and Social Care; Department for Transport; Department for Work and Pensions, which is coordinated by the National Institute for Health Research (NIHR, Ref: 198-1074). Funding has also been provided by the Economic and Social Research Council (ESRC). |
| **Ethics** | https://www.elsa-project.ac.uk/ethical-approval |

| | |
|---|---|
| **Website for Data Requests** | https://www.elsa-project.ac.uk/data-and-documentation |

| EXCEED: Extended Cohort for E-Health, Environment and DNA | |
|---|---|
| **Description of Study Population** (including citations and references if required) | EXCEED is a longitudinal population-based cohort which facilitates investigation of genetic, environmental and lifestyle-related determinants of a broad range of diseases and of multiple morbidity through data collected at baseline and via electronic healthcare record linkage. Recruitment has taken place in Leicester, Leicestershire and Rutland since 2013 and is ongoing, with 11,000 participants. Recruitment was widened to anyone over 18 years of age living in the Midlands in 2020. Participants provided a DNA sample, have consented to follow-up for up to 25 years through electronic health records and additional bespoke data collection is planned. Data available includes baseline demographics, anthropometry, spirometry, lifestyle factors (smoking and alcohol use) and longitudinal health information from primary care records, with additional linkage to other EHR datasets planned. Patients have consented to be contacted for recall-by-genotype and recall-by-phenotype sub-studies. Further details about the study can be accessed in the Cohort Profile Paper[1], with additional information about our COVID-19 Focus available as a Data Note Paper[2]<br><br>[1]Catherine John, Nicola F Reeve, Robert C Free, […] Edward J Hollox, Louise V Wain, Martin D Tobin. Cohort Profile: Extended Cohort for E-health, Environment and DNA (EXCEED). International Journal of Epidemiology, Volume 48, Issue 3, June 2019, Pages 678–679j, https://doi.org/10.1093/ije/dyz073<br><br>[2]Lee PH, Guyatt AL, John C et al. Extended Cohort for E-health, Environment and DNA (EXCEED) COVID-19 focus [version 1; peer review: awaiting peer review]. Wellcome Open Res 2021, 6:349, https://doi.org/10.12688/wellcomeopenres.17437.1 |

| | |
|---|---|
| **Acknowledgements** | EXCEED is funded by the University of Leicester, the NIHR Leicester Respiratory Biomedical Research Centre, the NIHR Clinical Research Network East Midlands, the Medical Research Council (grant G0902313) the Wellcome Trust (grant 202849) and HDR UK BREATHE- Health Data Research Hub for Respiratory Health (grant MC-PC_19004). |
| **Ethics** | The study is led by the University of Leicester, in partnership with University Hospitals of Leicester NHS Trust and in collaboration with Leicestershire Partnership NHS Trust, local general practices and smoking cessation services. Ethical approval for the study was obtained from the Leicester Central Research Ethics Committee (13/EM/0226). |
| **Website for Data Requests** | www.exceed.org.uk/research |

| Generation Scotland | |
|---|---|
| **Description of Study Population** (including citations and references if required) | The Generation Scotland Scottish Family Health Study has 24,000 adult volunteers recruited in Scotland 2006-2011, with consent for linkage to medical records and recontact for further studies [1].<br><br>www.ncbi.nlm.nih.gov/pubmed/?term=22786799<br><br>Smith, B. H., Campbell, A., Linksted, P., Fitzpatrick, B., Jackson, C., Kerr, S. M., … Morris, A. D. (2013). Cohort Profile: Generation Scotland: Scottish Family Health Study (GS:SFHS). The study, its participants and their potential for genetic research on health and illness. International Journal of Epidemiology, 42(3), 689-700. http://doi.org/10.1093/ije/dys084 |
| **Acknowledgements** | Generation Scotland received core support from the Chief Scientist Office of the Scottish Government Health Directorates [CZD/16/6] and the Scottish Funding Council |

| | |
|---|---|
| | [HR03006] and is currently supported by the Wellcome Trust [216767/Z/19/Z]. |
| **Ethics** | Generation Scotland obtained Research Tissue Bank approval from the East of Scotland Research Ethics Service (on behalf of NHS Scotland). Reference number 20/ES/0021. |
| **Website for Data Requests** | www.generationscotland.org |

| GLAD: Genetic Links to Anxiety and Depression Study dataset including the Eating Disorders Genetics Initiative (EDGI) and Covid-19 Psychiatry and Neurological Genetics Study (COPING) study | |
|---|---|
| **Description of Study Population** (including citations and references if required) | The Covid-19 Psychiatry and Neurological Genetics Study (COPING) was set up as a COVID-19 specific study investigating the mental health impact of individuals living in the UK (N=30,450). COPING participants were recruited from the existing GLAD, EDGI and NIHR Bioresource studies to provide pandemic relevant data. – for full details, see below – Young et al. (2021).<br><br>The Genetic Links to Anxiety and Depression (GLAD) Study is an NIHR Bioresource funded project assessing the genetic and environmental links to anxiety and depression (N=30,318) [1].<br><br>The Eating Disorders Genetics Initiative (EDGI) study is an NIHR Bioresource funded project assessing the genetic and environmental links to all eating disorders (N=3973).<br><br>GLAD –<br><br>[1]Davies, M. R., Kalsi, G., Armour, C., Jones, I. R., McIntosh, A. M., Smith, D. J., ... & Breen, G. (2019). The Genetic Links to Anxiety and Depression (GLAD) Study: Online recruitment into the largest recontactable study of depression and anxiety. *Behaviour Research and Therapy*, *123*, 103503. |

| | |
|---|---|
| | COPING –<br><br>Young, K. S., Purves, K. L., Hübel, C., Davies, M. R., Thompson, K. N., Bristow, S., ... & Breen, G. (2021). Depression, anxiety and PTSD symptoms before and during the COVID-19 pandemic in the UK.<br><br>EDGI –<br><br>N/A |
| **Acknowledgements** | The NIHR BioResource Centre Maudsley and the NIHR BioResource should be acknowledged in all publications resulting from any study that we have supported. Please use the following wording:<br><br>We thank the [delete as appropriate] GLAD/EDGI/COPING and [always keep] NIHR BioResource volunteers for their participation, and gratefully acknowledge NIHR BioResource centres, NHS Trusts and staff for their contribution. We thank the National Institute for Health Research, NHS Blood and Transplant, and Health Data Research UK as part of the Digital Innovation Hub Programme. This work was supported by the National Institute of Health Research (NIHR) BioResource Centre Maudsley and is part-funded by the National Institute for Health Research (NIHR) Biomedical Research Centre at South London and Maudsley NHS Foundation Trust and King’s College London. Patient and public involvement groups and services were provided by the NIHR KCL-Maudsley Biomedical Research Centre. The views expressed are those of the author(s) and not necessarily those of the NHS, the NIHR or the Department of Health and Social Care.<br><br>Any publication that makes substantial use of the data/participants provided by the GLAD Study should cite the following authors.<br><br>NIHR BioResource Consortium<br><br>Prof Gerome Breen |

| | |
|---|---|
| | Prof Thalia Eley<br><br>Prof Matthew Hotopf<br><br>Prof Cherie Armour<br><br>Prof Andrew McIntosh<br><br>Prof Ian R Jones<br><br>Dr Gursharan Kalsi<br><br>Dr Molly Davies<br><br>Dr Alish Palmos<br><br>Dr Christopher Huebel<br><br>Henry Rogers<br><br>Dina Monssen<br><br>Monika McAtarsney-Kovacs<br><br>Shannon Bristow<br><br>Ian Marsh<br><br>Kiran Glen<br><br>Chelsea Mika Malouf<br><br>Saakshi Kakar<br><br>Emily Kelly<br><br>Steven Bright<br><br>Affiliations: NIHR BioResource Centre Maudsley, King's College London, Denmark Hill, De Crespigny Park, London SE5 8AF<br><br>Centre for Social, Developmental and Psychiatric Genetics, IoPPN, King's College London, Denmark Hill, De Crespigny Park, London SE5 8AF<br><br>Any publication that makes substantial use of the data/participants provided by the EDGI Study should cite the following authors. |

| | |
|---|---|
| | NIHR BioResource Consortium<br><br>Prof Gerome Breen<br><br>Prof Thalia Eley<br><br>Prof Janet Treasure<br><br>Dr Moritz Herle<br><br>Dr Gursharan Kalsi<br><br>Dr Molly Davies<br><br>Dr Christopher Huebel<br><br>Dr Alish Palmos<br><br>Henry Rogers<br><br>Dina Monssen<br><br>Monika McAtarsney-Kovacs<br><br>Shannon Bristow<br><br>Ian Marsh<br><br>Kiran Glen<br><br>Chelsea Mika Malouf<br><br>Helena Davies<br><br>Emily Kelly<br><br>Saakshi Kakar<br><br>Affiliations: NIHR BioResource Centre Maudsley, King's College London, Denmark Hill, De Crespigny Park, London SE5 8AF<br><br>Centre for Social, Developmental and Psychiatric Genetics, IoPPN, King's College London, Denmark Hill, De Crespigny Park, London SE5 8AF |
| **Ethics** | GLAD – 18/LO/1218 – London Fulham REC<br><br>EDGI – 19/LO/1254 – London Fulham REC<br><br>COPING – 20/SW/0078 – South West Central Bristol REC |

| | |
|---|---|
| **Website for Data Requests** | https://gladstudy.org.uk (changing Autumn 2022)<br><br>https://edgiuk.org |

| MCS: Millennium Cohort Study | |
|---|---|
| **Description of Study Population** (including citations and references if required) | The Millennium Cohort Study (MCS) is following the lives of young people born across England, Scotland, Wales and Northern Ireland in 2000-02. The study began with an original sample of 18,818 cohort members. The study is designed and led by the Centre for Longitudinal Studies (CLS) at University College London.<br><br>The broad aim of the study is to examine the impact that circumstances and experiences at one stage of life have on outcomes and achievements in later life. Since the baseline survey at age 9 months, there have been six major 'sweeps' at ages 3, 5, 7, 11, 14 and 17. The next sweep, at age 22, is currently under development.<br><br>Data have been collected from a number of different sources, including the cohort members and their parents and teachers. The data have been collected in a variety of ways, including via paper and electronic questionnaires, biological samples, physical measurements, tests of ability, and linked educational attainment and health records.<br><br>The information collected forms a high quality data resource for scientific investigations across a full range of domains of individuals' lives and across different points in time in them. The study has been designed to ensure comparability with other major cohort studies both in the UK and internationally and to permit the examination of links between social change and the changing experiences of different cohorts.<br><br>https://www.llcsjournal.org/index.php/llcs/article/view/410/0<br><br>https://academic.oup.com/ije/article/43/6/1719/703283 |
| **Acknowledgements** | MCS is core-funded by the ESRC and co-funded by a consortium of government departments. |

| | |
|---|---|
| **Ethics** | Ethics approval has been obtained for each follow-up from an NHS Research Ethics Committee (REC). In addition, separate REC approval is in place to cover the ongoing activities of the study in between major sweeps of data collection (i.e. Keeping in touch with and tracing cohort Members; cleaning, documenting and providing access to the data for research and linking data from administrative sources to survey data to increase the utility of the data for research. |
| **Website for Data Requests** | https://cls.ucl.ac.uk/cls-studies/mcs/ |

| NCDS: National Child Development Study | |
| --- | --- |
| **Description of Study Population** (including citations and references if required) | The National Child Development Study (NCDS) is a continuing longitudinal study that seeks to follow the lives of all those living in Great Britain who were born in one particular week in 1958. Conducted by the Centre for Longitudinal Studies , the aim of the study is to improve understanding of the factors affecting human development over the whole lifespan. It collects information on physical and educational development, economic circumstances, employment, family life, health behaviour, wellbeing, social participation and attitudes.<br><br>The broad aim of the study is to examine the impact that circumstances and experiences at one stage of life have on outcomes and achievements in later life. Since the birth survey in 1958, there have been ten ‘sweeps’ of all cohort members at ages 7, 11, 16, 23, 33, 42, 44/5 (a biomedical collection) 46, 50 and most recently at 55. The Age 62 Sweep is currently in the field (2022).<br><br>Data have been collected from a number of different sources, including the midwife present at birth, parents of the cohort members, teachers, doctors and the cohort members themselves. The data have been collected in a variety of ways, including via paper and electronic questionnaires, clinical records, medical examinations, biological samples, physical measurements, tests of ability and educational assessments.<br><br>The information collected forms a high quality data resource for scientific investigations across a full range of domains of individuals’ lives and across different points in time in them. The study has been designed to ensure comparability with other major cohort studies and to permit the examination of links between social change and the changing experiences of different cohorts.<br><br>https://cls.ucl.ac.uk/cls-studies/1958-national-child-development-study/ |

| | |
|---|---|
| **Acknowledgements** | NCDS is core-funded by the ESRC. |
| **Ethics** | Ethics approval has been obtained for each follow-up from an NHS Research Ethics Committee (REC) since 2000. In addition, separate REC approval is in place to cover the ongoing activities of the study in between major sweeps of data collection (i.e. Keeping in touch with and tracing cohort members; cleaning, documenting and providing access to the data for research; and linking data from administrative sources to survey data to increase the utility of the data for research). |
| **Website for Data Requests** | https://cls.ucl.ac.uk/cls-studies/ncds/ |

| **Next Steps** | |
|---|---|
| **Description of Study Population** (including citations and references if required) | Next Steps (previously known as the Longitudinal Study of Young People in England (LSYPE1)) is a major longitudinal study that follows the lives of around 16,000 people born in 1989-90. The first seven sweeps of the study (2004-2010) were funded and managed by the Department for Education and mainly focused on the educational and early labour market experiences of young people.<br><br>The study began in 2004 and included young people in Year 9 who attended state and independent schools in England. Following the initial survey at age 13-14, the cohort members were interviewed every year until 2010.<br><br>In 2013 the management of Next Steps was transferred to the Centre for Longitudinal Studies (CLS) at the IOE, UCL's Faculty of Education and Society. The first sweep conducted by CLS aimed to find out how the lives of the cohort members had turned out at age 25. It maintained the strong focus on education, but the content was broadened to become a more multi-disciplinary research resource.<br><br>The Age 32 Sweep is currently in the field (2022). |

| | |
|---|---|
| | https://doc.ukdataservice.ac.uk/doc/5545/mrdoc/pdf/next_steps_userguide_to_the_redeposit_of_sweeps_1to7_may2020.pdf<br><br>https://doc.ukdataservice.ac.uk/doc/5545/mrdoc/pdf/nextsteps_age25_survey_user_guide_v3.pdf<br><br>https://cls.ucl.ac.uk/cls-studies/next-steps/ |
| **Acknowledgements** | Next Steps now is core-funded by the ESRC. |
| **Ethics** | Ethics approval is obtained for each follow-up from an NHS Research Ethics Committee (REC). In addition, separate REC approval is in place to cover the ongoing activities of the study in between major sweeps of data collection (i.e. keeping in touch with and tracing cohort members; cleaning, documenting and providing access to the data for research; and linking data from administrative sources to survey data to increase the utility of the data for research). |
| **Website for Data Requests** | https://cls.ucl.ac.uk/cls-studies/next-steps/ |

| NICOLA: Northern Ireland Cohort for the Longitudinal Study of Ageing | |
|---|---|
| **Description of Study Population** (including citations and references if required) | NICOLA is a large-scale longitudinal study in Northern Ireland designed to investigate ageing.<br><br>Set up in 2013 the study visited households where at least one member was $\geq$50 years old and in its first wave recruited a representative sample of 8478 men and women living in private residential accommodation in Northern Ireland. |

| | |
|---|---|
| | Wave 1 Data collection involved four components; a computer assisted face-to-face home interview, a self-completion questionnaire, a health assessment (during which blood and urine samples were collected) and subsequently a dietary questionnaire.<br><br>The participants who consented to follow up for Wave 2 were asked to complete a 2nd home survey and self-completion questionnaire approximately 3.5 years later. From these participants 5925 did not opt-out of additional linkage and were therefore in addition mailed a Covid questionnaire (3149 completed)<br><br>Neville CE, Cruise SM; Burns F. (2019). The Northern Ireland Cohort for the Longitudinal Study of Ageing (NICOLA). In Gu D, Dupre M. (Ed.), Encyclopedia of Gerontology and Population Aging Cham: Springer. |
| **Acknowledgements** | We are grateful to all the participants of the NICOLA Study, and the whole NICOLA team.<br><br>The study has received funding from The Atlantic Philanthropies, the Economic and Social Research Council, the UKCRC Centre of Excellence for Public Health Northern Ireland, the Centre for Ageing Research and Development in Ireland, the Office of the First Minister and Deputy First Minister, HSC Research and Development Division of the Public Health Agency, the Wellcome Trust/Wolfson Foundation and Queen's University Belfast which provide core financial support for NICOLA. The authors alone are responsible for the interpretation of the data and any views or opinions presented are solely those of the authors and do not necessarily represent those of the NICOLA Study team. |
| **Ethics** | Ethical approval for NICOLA was obtained from the School of Medicine, Dentistry and Biomedical Sciences Ethics Committee, Queen's University Belfast. |

| | |
|---|---|
| **Website for Data Requests** | https://www.qub.ac.uk/sites/NICOLA/InformationforResearchers/ |

| NIHR BioResource: National Institute of Health Research BioResource COVID-19 Psychiatry and Neurological Genetics (COPING) Study | |
|---|---|
| **Description of Study Population** (including citations and references if required) | The NIHR BioResource is a recallable resource of over 200,000 volunteers from the general population, and patients with rare and common diseases. Participants provide information about their health and lifestyle, together with biological samples, including DNA, and consent for access to their health records and for re-contact. The BioResource is one of four key infrastructures supporting population level genomic projects in the UK Life Science Industrial Strategy. Key unique features of the NIHR BioResource are its focus on recall of participants for experimental medicine studies by genotype and/or phenotype, and the inclusion of both healthy volunteers and patients with common and rare diseases. Participants represented in UK LLC are respondents to an online recall study, the Covid-19 Psychiatry and Neurological Genetics (COPING) study[1].<br><br>[1]https://www.maudsleybrc.nihr.ac.uk/posts/2020/may/covid-19-psychiatry-and-neurological-genetics-coping-study/ |
| **Acknowledgements** | We thank NIHR BioResource volunteers for their participation, and gratefully acknowledge NIHR BioResource centres, NHS Trusts and staff for their contribution. We thank the National Institute for Health and Care Research, NHS Blood and Transplant, and Health Data Research UK as part of the Digital Innovation Hub Programme. The views expressed are those of the author(s) and not necessarily those of the NHS, the NIHR or the Department of Health and Social Care. |
| **Ethics** | Research Tissue Bank (REC REF: 17/EE/0025). |
| **Website for Data Requests** | https://bioresource.nihr.ac.uk/using-our-bioresource/academic-and-clinical-researchers/apply-for-bioresource-data/ |

| TRACK-COVID Study |
|---|

| | |
|---|---|
| **Description of Study Population** (including citations and references if required) | The study employed a two-stage design. During the first stage of this effort, ~90,000 individuals previously recruited into the INTERVAL[1], COMPARE[2] and STRIDES studies (i.e., National Blood Donor Studies[3]) were invited via email to participate and to provide Covid-19-related information using an online questionnaire. During the second stage, participants were asked to provide self-collected finger-prick capillary blood sample every 6 weeks over a period of 18 months.<br><br>[1]Di Angelantonio E, Thompson SG, Kaptoge SK, Moore C, Walker M, Armitage J, Ouwehand WH, Roberts DJ, Danesh J, INTERVAL Trial Group. Efficiency and safety of varying the frequency of whole blood donation (INTERVAL): a randomised trial of 45 000 donors. Lancet. 2017 Nov 25;390(10110):2360-2371.<br><br>[2]Bell S, Sweeting M, Ramond A, Chung R, Kaptoge S, Walker M, Bolton T, Sambrook J, Moore C, McMahon A, Fahle S, Cullen D, Mehenny S, Wood AM, Armitage J, Ouwehand WO, Miflin G, Roberts DJ, Danesh J, Di Angelantonio E, COMPARE Study Group. Comparison of four methods to measure haemoglobin concentrations in whole blood donors (COMPARE): A diagnostic accuracy study. Transfus Med. 2020 Dec 20.<br><br>[3]http://www.donorhealth-btru.nihr.ac.uk/ |
| **Acknowledgements** | The TRACK-COVID study recruited participants from the INTERVAL trial and the COMPARE study and the academic coordinating centre would like to thank blood donor centre staff and blood donors for their participation. The centre was supported by core funding from the: National Institute for Health and Care Research (NIHR) Blood and Transplant Research Unit (BTRU) in Donor Health and Genomics (NIHR BTRU-2014-10024), NIHR BTRU in Donor Health and Behaviour (NIHR203337), British Heart Foundation (RG/13/13/30194; RG/18/13/33946) and NIHR Cambridge Biomedical Research Centre (BRC-1215-20014) [*].<br><br>We thank NIHR BioResource volunteers for their participation, and gratefully acknowledge NIHR BioResource centres, NHS Trusts and staff for their contribution. We thank the NIHR, NHS |

| | |
|---|---|
| | Blood and Transplant (NHSBT) and Health Data Research (HDR) UK as part of the Digital Innovation Hub Programme.<br><br>Participants in the COMPARE study were recruited with the active collaboration of NHSBT England (www.nhsbt.nhs.uk). Funding was provided by NHSBT and the NIHR BTRU in Donor Health and Genomics (NIHR BTRU-2014-10024). DNA extraction and genotyping were co-funded by the NIHR BTRU and the NIHR BioResource (http://bioresource.nihr.ac.uk). The academic coordinating centre for COMPARE was supported by core funding from the: NIHR BTRU in Donor Health and Genomics, UK MRC (MR/L003120/1), BHF (RG/13/13/30194; RG/18/13/33946) and NIHR Cambridge BRC (BRC-1215-20014) [*]. A complete list of the investigators and contributors to the COMPARE study is provided in reference [**]. The academic coordinating centre would like to thank blood donor centre staff and blood donors for participating in the COMPARE study.<br><br>Participants in the INTERVAL randomised controlled trial were recruited with the active collaboration of NHSBT, which has supported field work and other elements of the trial. DNA extraction and genotyping were co-funded by the NIHR, the NIHR BioResource and the NIHR Cambridge BRC (BRC-1215-20014) [*]. The academic coordinating centre for INTERVAL was supported by core funding from the: NIHR BTRU in Donor Health and Genomics (NIHR BTRU-2014-10024), UK MRC (MR/L003120/1), BHF (SP/09/002; RG/13/13/30194; RG/18/13/33946) and NIHR Cambridge BRC (BRC-1215-20014) [*]. A complete list of the investigators and contributors to the INTERVAL trial is provided in reference [***]. The academic coordinating centre would like to thank blood donor centre staff and blood donors for participating in the INTERVAL trial.<br><br>This work was also supported by HDR UK, which is funded by the UK Medical Research Council, Engineering and Physical Sciences Research Council, Economic and Social Research Council, Department of Health and Social Care (England), Chief Scientist Office of the Scottish Government Health and Social Care Directorates, Health and Social Care Research and Development Division (Welsh Government), Public Health |

| | |
|---|---|
| | Agency (Northern Ireland), British Heart Foundation and Wellcome.<br><br>We thank Leeds Teaching Hospitals NHS Foundation Trust for their contribution to the SARS-COV-2 antibody analysis.<br><br>*The views expressed are those of the author(s) and not necessarily those of the NIHR, NHSBT or the Department of Health and Social Care.<br><br>**Bell S, Sweeting M, Ramond A, Chung R, Kaptoge S, Walker M, Bolton T, Sambrook J, Moore C, McMahon A, Fahle S, Cullen D, Mehenny S, Wood AM, Armitage J, Ouwehand WO, Miflin G, Roberts DJ, Danesh J, Di Angelantonio E, COMPARE Study Group. Comparison of four methods to measure haemoglobin concentrations in whole blood donors (COMPARE): A diagnostic accuracy study. Transfus Med. 2020 Dec 20.<br><br>***Di Angelantonio E, Thompson SG, Kaptoge SK, Moore C, Walker M, Armitage J, Ouwehand WH, Roberts DJ, Danesh J, INTERVAL Trial Group. Efficiency and safety of varying the frequency of whole blood donation (INTERVAL): a randomised trial of 45 000 donors. Lancet. 2017 Nov 25;390(10110):2360-2371. |
| **Ethics** | 20/EM/0121 - East Midlands - Nottingham 2 Research Ethics Committee |
| **Website for Data Requests** | http://www.donorhealth-btru.nihr.ac.uk/ |

| TwinsUK | |
|---|---|
| **Description of Study Population** (including citations and references if required) | TwinsUK is the largest adult twin registry in the UK and the most clinically detailed in the world. The national, population-based study was founded in 1992 and aims to investigate the genetic and environmental basis of a range of complex diseases and conditions. TwinsUK currently consists of over 15,700 volunteer adult twins (both monozygotic and dizygotic) who are between 18 to 104 years of age from around the UK (mean |

| | |
|---|---|
| | age 59)[1]. The cohort is predominantly female, and disease prevalence is broadly reflective of the UK population. Over 700,000 biological samples and extensive phenotypes have been collected longitudinally over 30 years.<br><br>[1]Verdi S, Abbasian G, Bowyer RCE, et al.: TwinsUK: The UK Adult Twin Registry Update. Twin Res Hum Genet. 2019; 22(6): 523–529. |
| **Acknowledgements** | We thank TwinsUK members for their participation and the TwinsUK operations team for coordinating and undertaking twin clinic visits and data and sample collections.<br><br>TwinsUK is funded by the Wellcome Trust, Medical Research Council, Versus Arthritis, European Union Horizon 2020, Chronic Disease Research Foundation (CDRF), Zoe Ltd, the National Institute for Health and Care Research (NIHR) Clinical Research Network (CRN) and Biomedical Research Centre based at Guy's and St Thomas' NHS Foundation Trust in partnership with King's College London. |
| **Ethics** | All collections of TwinsUK data have received ethical approval associated with TwinsUK Biobank (19/NW/0187), TwinsUK (EC04/015) or Healthy Ageing Twin Study (H.A.T.S) (07/H0802/84) from NHS Research Ethics Committees. Linkage to health and environmental records is also covered by approval from the Health Research Authority (19/CAG/0223). |
| **Website for Data Requests** | https://twinsuk.ac.uk/resources-for-researchers/access-our-data/ |

| **Understanding Society – the UK Household Longitudinal Study** | |
|---|---|
| **Description of Study Population** (including citations and references if required) | Understanding Society, the UK Household Longitudinal Study, is a longitudinal survey of the members of ~40,000 households (at Wave 1, 2009-10) in the United Kingdom. The survey sample consists of a large General Population Sample (~26,000 |

| | |
|---|---|
| | households) plus three other components: the Ethnic Minority Boost Sample (~4,000 households), the former British Household Panel Survey sample (~8,000 households) and the Immigrant and Ethnic Minority Boost Sample (~2,900 households, added at Wave 6). Household and individual interviews are conducted annually. The study is multi-topic and multi-purpose.<br><br>From April 2020 to September 2021, participants from the main Understanding Society sample were asked to complete nine short web-surveys (with a telephone option in some months). The COVID-19 study covered the changing impact of the pandemic on the welfare of UK individuals, families and wider communities. ~18,000 individuals provided a full or partial interview at Wave 1 (April 2020).<br><br>At Wave 8 of the COVID-19 study, 8477 participants provided consent to link their survey data to administrative health records. |
| **Acknowledgements** | Understanding Society is an initiative funded by the Economic and Social Research Council and various Government Departments, with scientific leadership by the Institute for Social and Economic Research, University of Essex, and survey delivery by NatCen Social Research and Kantar Public<br><br>The COVID-19 study (2020-2021) was funded by the Economic and Social Research Council and the Health Foundation. Serology testing was funded by the COVID-19 Longitudinal Health and Wealth – National Core Study. Fieldwork for the web survey was carried out by Ipsos MORI and for the telephone survey by Kantar. |
| **Ethics** | The University of Essex Ethics Committee has approved all data collection on Understanding Society main study, COVID-19 surveys and innovation panel waves, including asking consent for all data linkages except to health records. |

| | |
|---|---|
| | Approval for asking consent for health record linkage and for the collection of blood and subsequent serology testing in the March 2021 wave of the COVID-19 study was obtained from London – City & East Research Ethics Committee (21/HRA/0644). |
| **Website for Data Requests** | https://ukllc.ac.uk or https://ukdataservice.ac.uk |
